# A combined polygenic score of 21,293 rare and 22 common variants significantly improves diabetes diagnosis based on hemoglobin A1C levels

**DOI:** 10.1101/2021.11.04.21265868

**Authors:** Peter Dornbos, Ryan Koesterer, Andrew Ruttenburg, Joanne B. Cole, AMP-T2D-GENES Consortia, Aaron Leong, James B. Meigs, Jose C. Florez, Jerome I. Rotter, Miriam S. Udler, Jason Flannick

## Abstract

Polygenic scores (PS), constructed from the combined effects of many genetic variants^1^, have been shown to predict risk or treatment strategies for certain common diseases^2–6^. As most PS to date are based on common variants^7^, the benefit of adding rare variation to PS remains largely unknown and methodically challenging. We developed and validated a method for constructing a rare variant PS and applied it to a previously identified clinical scenario, in which genetic variants modify the hemoglobin A1C (HbA1C) threshold recommended for type 2 diabetes (T2D) diagnosis^6, 8–10^. The resultant rare variant PS is highly polygenic (21,293 variants across 144 genes), depends on ultra-rare variants (72.7% of variants observed in <3 people), and identifies significantly more undiagnosed T2D cases than expected by chance (OR=2.71, *p*=1.51×10^-6^). A model combining the rare variant PS with a previously published common variant PS^6^ is expected to identify 4.9M misdiagnosed T2D cases in the USA, nearly 1.5-fold more than the common variant PS alone. These results provide a method for constructing complex trait PS from rare variants and suggest that rare variants will augment common variants in precision medicine approaches for common disease.

## Main

HbA1C, which measures average blood glucose levels over 2-3 months^12^, is a widely used T2D diagnostic biomarker. Meeting the HbA1C diagnostic threshold of 6.5%^13^ – as opposed to a milder clinical diagnosis of pre-diabetes for individuals near the threshold^14^ – has both insurance and treatment implications, as the therapeutic armamentarium is much larger once a T2D diagnosis is made^15, 16^. An individual’s HbA1C is influenced by common genetic variants that affect both pathways central to glycemic control^6, 9, 10^ and pathways that influence erythrocytic properties such as cell lifespan^17^. Erythrocytic variants do not contribute to risk of T2D and can in fact cause diabetes misdiagnoses by altering the relationship between measured HbA1C and true blood glucose levels^6, 9, 10, 18, 19^. Accounting for the effects of common erythrocytic variants through a PS^6^ identifies a substantial number of undiagnosed T2D cases in the population – most notably, roughly 11% of African-Americans carry a *G6PD* variant (rs1050828)^6, 20^ that shortens the mean erythrocyte lifespan and may cause ∼0.65M African-American individuals with T2D to be undiagnosed^6, 10^.

The identification of common variants that affect HbA1C through erythrocytic pathways advances the public health goal of ensuring that all people across all genetic backgrounds are diagnosed accurately for diabetes. Most variants in a population, however, are rare^21^ and have unknown impacts on HbA1C or the T2D diagnostic threshold. Furthermore, while rare variants have been used to predict^22^ and diagnose^23^ Mendelian diseases, it remains unknown if they could meaningfully contribute to PS for complex traits such as T2D or HbA1C. Current methodologies for common variant PS, which rely on effect size estimates for variants from a “discovery” study^1, 2, 7^, do not obviously extend to incorporate rare variants, because (a) most individual rare variant effect sizes cannot be accurately estimated^24, 25^ and (b) most rare variants carried by a patient are absent from even large discovery studies^21^.

To include and evaluate the utility of rare coding variants in a PS for the HbA1C- based T2D diagnostic threshold, we analyzed whole exome sequence (WES) data in up to 87,735 individuals (**Supplementary Table 1**) from the UK biobank^26, 27^ (UKB; N≤45,650 Europeans [EU]) and AMP-T2D-GENES (N≤12,132 EU; 12,369 Hispanics [HS]; 6,234 African-Americans [AA]; 5,931 East Asians [EA]; 5,419 South Asians [SA])^28^. A single-variant association analysis across HbA1C and 23 other phenotypes (conducted to compare HbA1C association strengths to those for other metabolic phenotypes; **Supplementary Table 2**) produced 51 nonsynonymous coding variant HbA1C associations at the study-specific exome-wide significance threshold (*p*≤1.8×10^-8^ [**Methods**]; **Supplementary Figure 1**), the most of any non-lipid phenotype (**Figure 1a**). Most (N=46) of these associations were with common or low frequency variants (MAF ≥ 0.01), all were in known HbA1C GWAS loci (**Methods**), and all had smaller effect sizes than other known associations in their respective loci (**Methods**; **Supplementary Table 3**). Consequently, none of these variants significantly augmented the previous common variant HbA1C PS.

**Figure 1.**
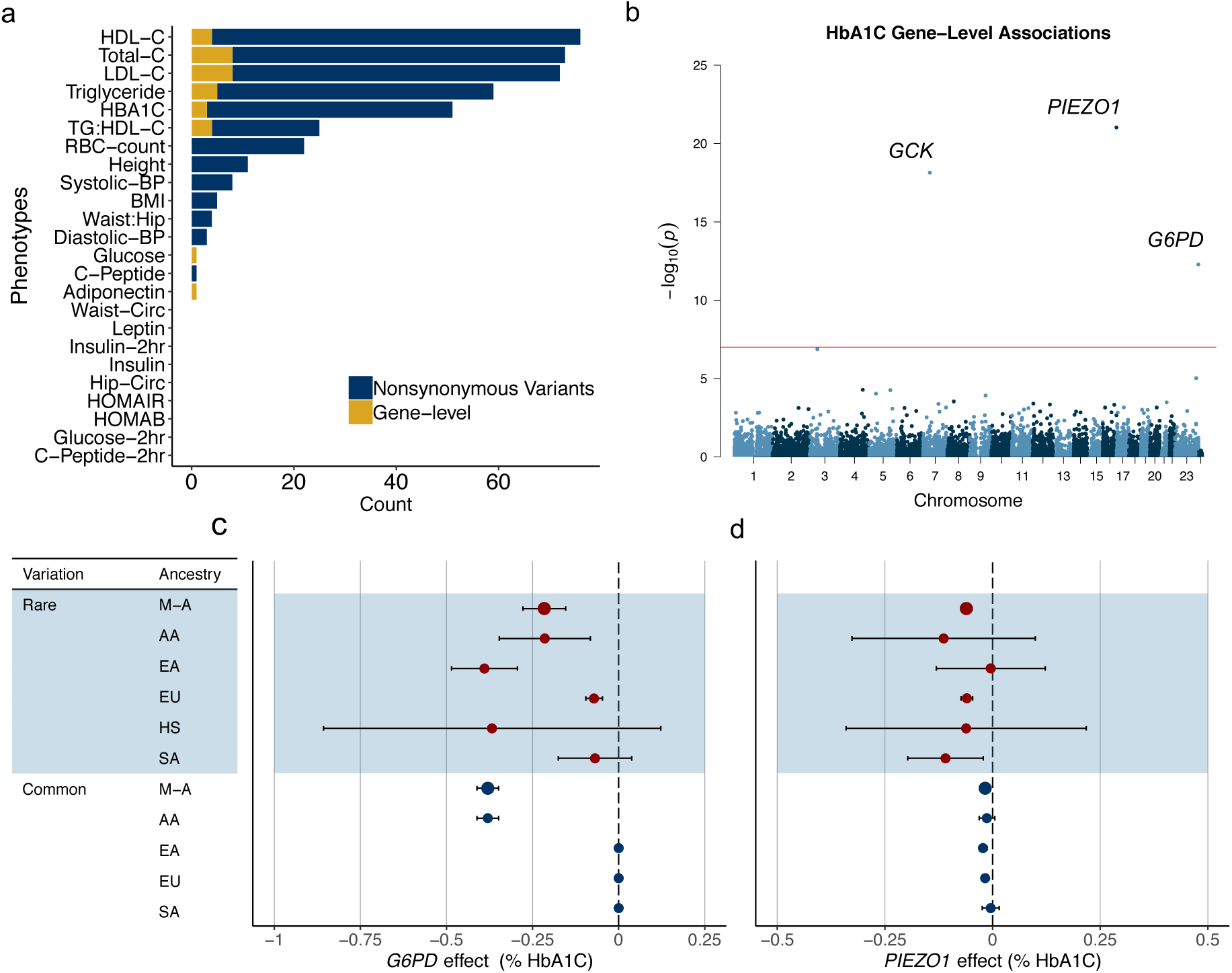
Rare variant associations for HbA1C are comparatively strong. **(a)** The number of exome-wide significant nonsynonymous variant associations (*p*≤1.8 10^-8^; yellow) and exome-wide significant gene-level associations (*p*≤1.0 10^-7^; blue) across 24 quantitative phenotypes. **(b)** Manhattan plot of all gene-level HbA1C associations. Those reaching exome-wide significance (*p* 1.0 10^-7^; red line) are labeled. **(c-d)** Effect sizes (percent glycated hemoglobin units) for rare variant gene-level associations and nearby common variant associations for **(c)** *G6PD* and **(d)** *PIEZO1*. Gene-level effects displayed are from the strongest associated variant mask. Abbreviations: AA indicates African-American, EA indicates East Asian, EU indicates European, HS indicates Hispanic, and SA indicates South Asian. M-A indicates meta-analysis across ancestries.

We next focused on variants too rare for individual analysis by conducting a gene-level analysis, in which we grouped variants into seven nested “masks” and conducted aggregate burden tests for each gene (**Methods**; **Supplementary Table 4**). Three HbA1C gene-level associations reached the study-specific exome-wide significance threshold (*p*≤1.0×10^-7^; **Figure 1b**), again more than any non-lipid phenotype (**Figure 1a**). Two of these three genes, *PIEZO1* (effect=-0.06% HbA1C; *p*=2.8×10^-23^) and *G6PD* (-0.22%; *p*=3.2×10^-10^), have known roles in erythrocyte function^29–34^. Both associations had smaller aggregate effect sizes than previously estimated for the common *G6PD* rs1050828 variant (-0.38%)^6^, but both had larger effect sizes than typical common variant HbA1C associations (∼0.024%)(**Figure 1c-d**; **Supplementary Table 5**)^6^. In fact, the *PIEZO1* gene-level association explained more HbA1C phenotype variance than a nearby GWAS association^6^ (**Supplementary Figure 2**), which was not due to LD between the signals (**Supplementary Table 6**; **Methods**) and which stands in contrast to the properties of rare variant gene-level associations previously reported for T2D^28^.

In light of the strength of the *PIEZO1* and *G6PD* associations (**Figure 1c-d**), the known erythrocytic roles of these genes, and previous work with common variants^6, 9^, we hypothesized that rare variants in *PIEZO1* and *G6PD* might alter the appropriate HbA1C diagnostic threshold for T2D. To test this hypothesis, we identified individuals in AMP-T2D-GENES (our “test sample”) who (a) had HbA1C levels below 6.5%, (b) had not taken antihyperglycemic medication, and (c) carried *PIEZO1* or *G6PD* variants with effect sizes (as estimated from our “discovery sample”, the UKB) sufficient to adjust their HbA1C levels above 6.5% (“reclassify” them). We then evaluated the proportion of “true” T2D cases (defined by non-HbA1C measurements; **Methods**) among the reclassified carriers and compared this to the proportion of true cases among individuals matched on cohort and HbA1C level (**Methods**). To validate this approach, we applied it to the published common variant HbA1C PS^6^ for four different ancestries (**Methods**; **Supplementary Figure 3**) and found that, in our test sample, the PS together reclassified 1.0% of individuals and a greater proportion of true cases than expected by chance (OR=3.42; *p*=3.3×10^-8^). We then used the approach to evaluate the variants in the *PIEZO1* and *G6PD* gene-level associations, assigning each variant an effect size equal to that measured for the aggregate association (**Methods**). We observed 0.2% of samples to be reclassified, including only marginally more true cases than expected by chance (OR=1.67; *p*=0.29). Thus, the gene-level *PIEZO1* and *G6PD* associations affect the HbA1C diagnostic threshold for fewer people and with less accuracy than does the common variant PS^6^.

To explore rare variants beyond those reaching exome-wide significance, we conducted two enrichment analyses. First, following previous HbA1C GWAS^6^, we observed the strongest gene-level HbA1C associations to be moderately enriched for rare variant erythrocyte count (RBC) associations (lowest *q*-value=0.04; **Methods**; **Supplementary Figure 4**). Second, following a previous T2D WES study^28^ we tested for the enrichment of gene-level HbA1C associations within gene sets defined from knockout mouse studies (**Methods**). We observed *p*<0.05 enrichments within 6 of 25 gene sets curated from glycemic effects in mice and 4 of 10 gene sets curated from erythrocytic effects in mice (**Figure 2a**; **Methods**). Moreover, the *p*<0.05 gene-level associations within the four enriched erythrocytic gene sets had effect sizes biased toward decreased HbA1C (16/23, binomial *p*=0.04; **Figure 2b**), consistent with expectations from the gene set annotation and previous observations in humans^17^. These results suggest that a substantial number of genes that do not reach exome-wide significance in our analysis harbor rare variants that not only affect HbA1C levels but do so independently of glycemia.

**Figure 2.**
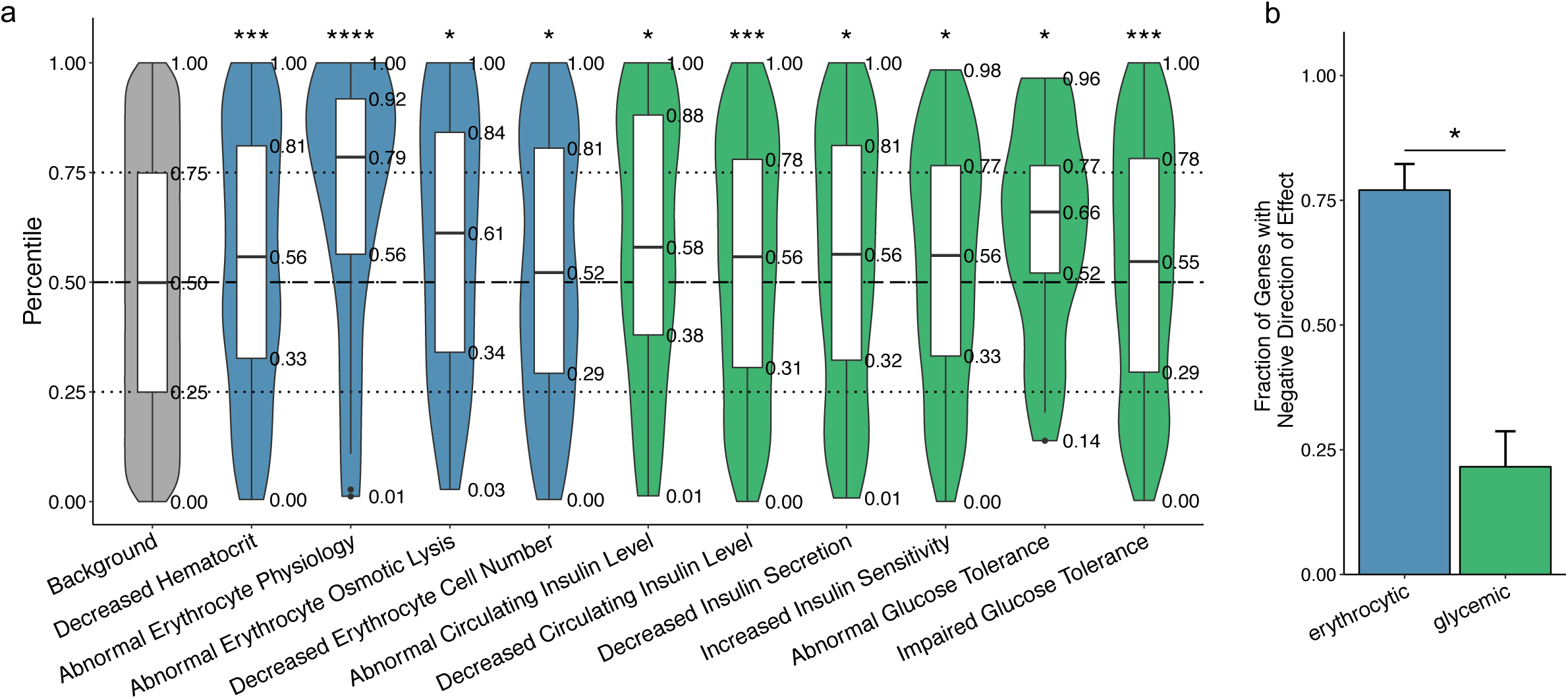
Rare variant gene-level HbA1C associations show enrichment for genes involved in glycemic control and erythrocytic pathways in mice. **(a)** Boxplots displaying the percentile of gene-level *p*-values (relative to matched genes; grey) for genes known to impact erythrocyte pathways (blue) and glycemic control (green) in mice. The horizontal dotted line at the 50^th^ percentile indicates the expected median percentile under the null distribution; we used a Wilcoxon rank sum test to assess significant deviation of percentiles from the null. **(b)** The fraction of genes with a negative effect on HbA1C levels among those (i) with HbA1C gene-level *p*≤0.05 and (ii) within a significantly enriched (*p*≤0.05) erythrocytic or glycemic gene set. We used a binomial test to assess deviation from the expected fraction of 50%. In each panel, asterisks indicate significance (*= ≤0.05; **= ≤5.0×10^-3^; ***=≤5.0×10^-4^; ****=≤5.0×10^-5^).

We next sought to augment the existing HbA1C common variant PS with this broader collection of rare variants. Existing PS methods^1–4, 35, 36^, however, are designed for common variants and do not extend easily to include many rare variants across many genes. Challenges include (a) selecting genes to include in the PS; (b) assigning weights to variants within the selected genes; and (c) assigning weights to rare variants not seen in the discovery sample. Rare coding variant PS also, however, present new opportunities: numerous annotations of gene function are publicly available to help select genes^37–41^, and estimates of functional effects are easier to incorporate into weights for coding variants than they are for non-coding variants^24, 42, 43^. We therefore developed a novel framework for a rare coding variant HbA1C PS that (a) includes genes based on their aggregate *p*-values and experimental annotations (*e.g.* from knockout mice; **Figure 3a**) and (b) assigns variant weights based on aggregate effect sizes for the masks that contain the variant (**Methods**; **Figure 3b**). We explored 12 potential models within this framework by varying the precise criteria used for gene selection and weight assignment (**Figure 3c**).

**Figure 3.**
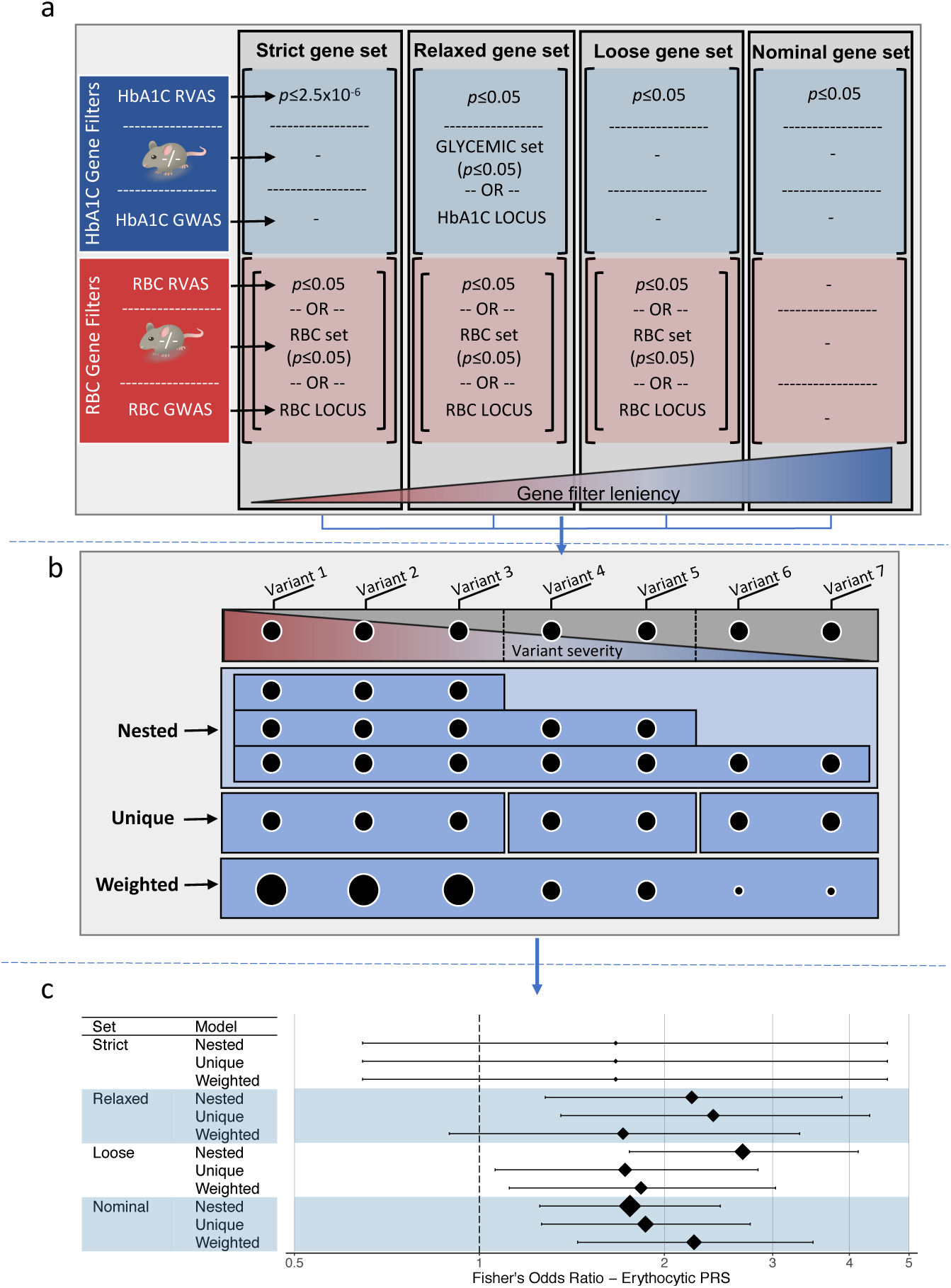
A framework for constructing polygenic scores that include rare variants. The framework consists of three steps: **(a)** choosing genes to include in the polygenic score, based on their association *p*-value and annotation; **(b)** defining weights for rare variants, based on the masks that include them and the aggregate effect sizes observed for the masks, and **(c)** testing the accuracy of the polygenic score by identifying T2D cases with adjusted HbA1C that crosses the diagnostic threshold (6.5% HbA1C) and comparing (via a Fisher’s exact test) the number of cases that cross the threshold to the number expected by chance. **(a)** We explored four methods for choosing genes, based on their strength of HbA1C association (blue boxes) and evidence of acting through erythrocytic pathways (red). “GLYCEMIC set” and “RBC set” indicate genes located within an enriched (at *p*≤0.05) glycemic or erythrocytic gene set (Figure 2). “HbA1C LOCUS” and “RBC LOCUS” indicate genes located within 125kb of a common variant HbA1C or RBC association. **(b)** We explored three methods for weighting variants: the aggregate effect size of the strictest mask that contained the variant (nested), the aggregate effect size of variants unique to the strictest mask that contained the variant (unique), or the aggregate effect size of a weighted burden test for the gene (weighted). **(c)** For each of the twelve resulting rare variant polygenic scores, we calculated odds ratios and 95% confidence intervals for the fraction of true T2D cases reclassified by the model as compared to the null expectation (controlled for HbA1C levels). The area of the diamond for each odds ratio is proportional to the total number of individuals in our test sample reclassified by the model.

Six (50%) of the tested PS models reclassified a significantly greater (OR>1, *p*≤4.1×10^-3^ [0.05/12 models]) proportion of true cases than expected by chance (**Figure 3c**). The most significant excess of reclassified true cases (OR=2.71, *p*=1.5×10^-6^; **Figure 3c**) was achieved by a model with a “loose” criterion for gene-level HbA1C association and a “nested” method for variant weights (**Methods**; **Figure 3a, 3b**). The “loose” criterion requires genes to achieve HbA1C rare variant *p*<0.05 and then, to filter for evidence of acting through erythrocytic pathways, includes only genes that either have a nominal (*p*<0.05) RBC rare variant association, are within 125kb of an RBC GWAS association, or are in an enriched mouse erythrocytic gene set. The “nested” method assigns each variant a weight equal to the aggregate HbA1C effect size (in the UKB discovery sample) of variants with annotations at least as severe as the variant (*i.e.* in the most severe nested mask containing the variant; **Methods**). This “loose, nested” model (**Methods**) contains 144 genes and 21,293 variants from our study (**Supplementary Table 7**), with 21,016 (98.7%) variants having MAF<0.005 and 15,473 (72.7%) observed in <3 people. Of individuals in our test sample, 20,927 (99.7%) carry a variant with non-zero weight in this model, including 13,573 (64.6%) who carry a variant absent from the discovery study (**Methods**). As a negative control, we replaced the erythrocyte gene filters in the model with filters for genes involved in glucose homeostasis (**Methods**) and found that the resultant model did not reclassify significantly more true cases than expected by chance (OR=1.38, *p*=0.16).

Next, we evaluated whether the “loose, nested” rare variant PS could augment the previous common variant HbA1C PS^6^ (**Supplementary Table 8**; **Methods**). A combined PS summing the ancestry-appropriate common variant PS with the rare variant PS (with additivity justified by the independence of variants in the rare and common variant PS; **Methods**, **Supplementary Figure 5**) produced a greater reclassification accuracy in our test sample (OR=4.4, *p*=8.0×10^-26^) than either the rare or common PS alone. To then calculate the number of people in the US population who might have a T2D diagnosis altered by this combined PS, we scaled the ancestry proportions in our test sample to the estimated ancestry proportions of the US population (**Methods**). This calculation suggested that the combined PS is expected to re-classify 4.9M US T2D cases, nearly 1.5-fold more than the common variant PS alone (3.4M) and 2-fold more than the rare variant PS alone (2.4M) (**Figure 4a**).

**Figure 4.**
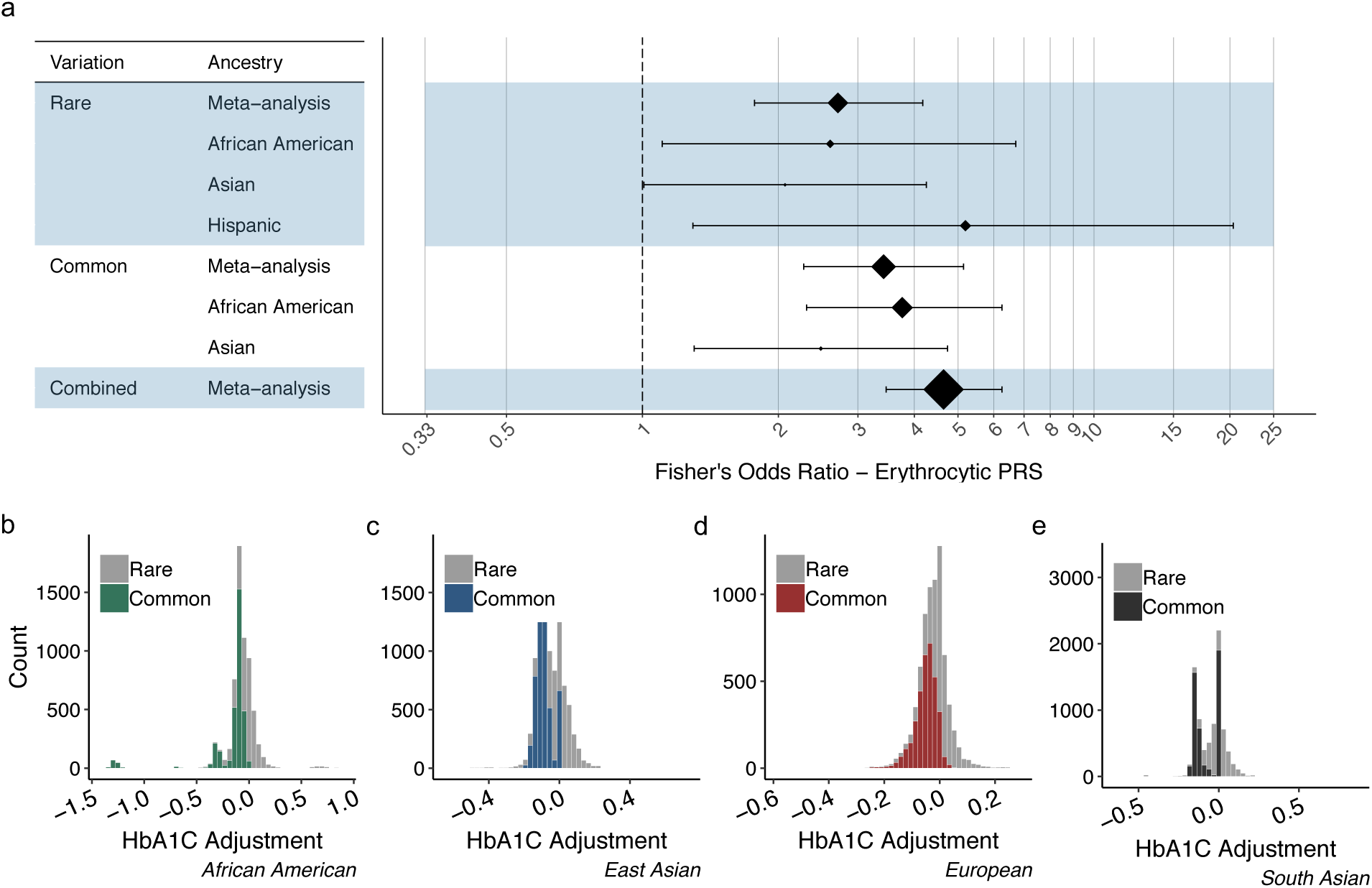
The accuracy and properties of rare and common variant polygenic scores. **(a)** The forest plot indicates Fisher’s exact test odds ratios and 95% confidence intervals (as in Figure 3c) for the best performing rare variant polygenic score (the ‘nested variant’ ‘loose’ model), a common variant polygenic score (based on a previous GWAS^6^), and a combined polygenic score. The area of each diamond is proportional to the estimated number of individuals in the US population that would be reclassified after scaling the ancestral proportions in our test sample to those estimated for the US (**Methods**). In the figure, “Asian” represents a meta-analysis of the South Asian and East Asian results; Europeans are not displayed due to insufficient data in our test sample. **(b-e)** Histograms display the distribution (in % HbA1C) of rare variant (gray) and common variant (colored) polygenic scores for each ancestry.

One reason that the rare variant PS compares favorably to the common variant PS in the US population is that a common variant PS does not exist for ancestries without GWAS effect size estimates (*e.g.* Hispanics^6^), while the rare variant PS – because variants included in it depend on molecular annotations rather than ancestry-specific effect sizes – is defined for all ancestries (**Figure 3a**, **4a**). While further work is needed to fully justify the use of fixed rare variant weights across ancestries, our initial results suggest that (a) the accuracy of the rare variant PS is comparable across all ancestries in our test sample (**Figure 4a**) and (b) gene-level effect sizes in the rare variant PS are more homogenous across ancestries than are the variant-level effect sizes in the common variant PS (λ=3.1 for common variant Q statistic *vs.* λ≤1.6 for gene-level Q statistic[s]; **Supplementary Figure 6**; **Methods**). This observation does not conflict with the known population-specificity of rare variants^21, 44^, as the rare variant PS is more variable across ancestries than the common variant PS in terms of the specific variants it includes: most (68%) of the variants in the rare variant PS are ancestry-specific, and the average number of rare variants carried by an individual varies substantially by ancestry (**Supplementary Table 9**).

Collectively, these results suggest that rare variants – despite a comparatively modest impact on complex traits^28, 45^ – may meaningfully contribute to genetic-based diagnostic strategies for complex disease. The properties of our rare variant PS are contrary, however, to some expectations that rare variants will be useful in genetic risk prediction primarily through population-specific large effects^21, 44, 46^. We indeed find that the average aggregate effect size for genes included in the rare variant model is larger than the average effect size of variants in the common variant model (0.066% ± 0.005% *vs*. 0.0349% ± 0.017% average absolute effect; **Supplementary Table 10**). However, the average PS adjustment across all individuals from the rare variant model (0.056% HbA1C) is weaker than that from the common variant model (0.10% HbA1C; **Supplementary Table 11**), because individuals tend to carry fewer variants in the rare variant PS than they do in the common variant PS (4.7 *vs.* 8.4) – even in spite of the much larger number of variants in the rare variant PS (21,293 *vs.* 22; **Supplementary Table 9**). The properties of the rare variant PS are thus natural consequences of the difference between properties of rare variants individually (large effect and ancestry specific) and their properties in aggregate (polygenic and relatively ancestry non-specific; **Figure 4b-e**).

Rare variant PS also complement common variant PS in other aspects. First, while both types of PS benefit by including suggestive (*p*<0.05) but not exome-wide significant associations, we show that incorporating gene annotations from model organisms (*e.g.* knockout mice) increases rare variant PS accuracy. Other gene annotations^47^, which are currently more readily available than noncoding variant annotations^48^, could naturally be used to further improve the model. As noncoding variant annotations do improve, we anticipate that extending rare variant PS to include noncoding variants could likewise improve PS accuracy^49^. A second property of the rare variant PS is that it does not require individual variant effect size estimates. It can thus incorporate variants private to an individual, and our data provide evidence that its variant weights can be held roughly constant across ancestries. While limited in a strict sense to HbA1C adjustments for diabetes diagnosis, in a broader sense – given the broad similarities in genetic architectures of many complex traits^50, 51^ – the current study is an initial step towards rare variant PS for complex traits in general. Our results suggest that rare variants will be a valuable addition to complex trait PS, albeit through polygenic effects that look quite distinct from those that might have been expected from early predictions of rare variants with a “high impact” on common disease^24, 52^.

## Methods

### AMP-T2D-GENES analysis

#### AMP-T2D-GENES cohort and phenotype information

The AMP-T2D-GENES dataset was the focus of a previous T2D exome sequence analysis^28^. It is comprised of samples from five ancestries (African American, East Asian, European, Hispanic, and South Asian) and six consortia: (a) T2D Genetic Exploration by Next-gen sequencing of multi-Ethnic Samples (T2D-GENES)^45^, (b) Slim Initiative Genomic Medicine for the Americas: T2D (SIGMA T2D)^53^, (c) Genetics of T2D (GoT2D)^45^, (d) Exome-sequencing Project (ESP)^21^, (e) Lundbeck Foundation Centre for Applied Medical Genomics in Personalized Disease Predictions, Prevention, and Care (LuCamp) study^54^, (f) Progress in Diabetes Genetics in Youth (ProDiGY)^55^. The dataset consists of sample genotypes, covariates (*e.g.* race, age, and sex), and phenotypes for up to 23 quantitative phenotypes: body mass index (BMI), diastolic blood pressure (Diastolic-BP), systolic blood pressure (Systolic-BP), total cholesterol (Total-C), triglyceride, low-density lipoprotein cholesterol (LDL-C), high-density lipoprotein cholesterol (HDL-C), waist-hip ratio (Waist:Hip), Height, hemoglobin A1C (HbA1C), triglyceride:HDL-C ratio (TG:HDL-C), hip circumference (Hip-Circ), waist circumference (Waist-Circ), fasting glucose (Glucose), fasting insulin (Insulin), homeostatic model assessment for insulin resistance (HOMAIR), homeostatic model assessment of β function (HOMAB), Adiponectin, glucose level post 2hr glucose tolerance test (Glucose-2hr), leptin, insulin level post 2hr glucose tolerance test (Insulin-2hr), C-peptide level (C-peptide), C-peptide level post 2hr glucose tolerance test (C-peptide-2hr). For glycemia-related phenotypes (HbA1C, Glucose, Insulin, HOMAIR, HOMAB, Adiponectin, Glucose-2hr, leptin, Insulin-2hr, C-peptide, and C-peptide-2hr), only T2D control subjects (see ‘**AMP-T2D-GENES type 2 diabetes case definition**’ methods) were included in association analyses. As not all samples have measurements for each phenotype, association sample sizes vary across the phenotypes (**Supplementary Table 1**). Cohort-level summary statistics for each phenotype are presented in **Supplementary Table 12**.

All individuals in the AMP-T2D-GENES study provided informed consent and all samples were approved for use at the respective institution^21, 28, 45, 53, 54^.

#### AMP-T2D-GENES phenotype transformations

Prior to association analyses, we transformed phenotypes to more closely follow a normal distribution. We first log-transformed phenotypes with skewed distributions (Insulin-2hr, Adiponectin, BMI, Insulin, Glucose-2hr, HbA1C, HDL-C, Hip-Circ, HOMAB, HOMAIR, TG:HDL-C, Systolic-BP, Triglyceride, and Leptin). We then adjusted all phenotypes for age, age^2^, and sex using residuals from linear regression^56^. Finally, we inverse-normal transformed the calculated residuals for each phenotype and multiplied the resulting values by the standard deviation of the original phenotype distribution (to scale back to the original units used for the phenotype measurements). We performed these transformations separately within each cohort of the AMP-T2D-GENES dataset.

#### AMP-T2D-GENES type 2 diabetes case definition

The AMP-T2D-GENES T2D case definition was based on previously described diagnostic thresholds^28^. Individuals were categorized as a T2D case if: (a) a 2-hour glucose measure was ≥1.1 mmol/L or (b) any two of the following suggestive measures were met:

(1) fasting glucose levels ≥
(2) HbA1C levels ≥ 5%
(3) there is a record of antihyperglycemic medication use.

When conducting our reclassification analysis for the HbA1C PS, we considered as true cases only those individuals who had criteria (1) or (3) satisfied – that is, we ignored the HbA1C based criterion. All individuals not categorized as T2D cases were considered controls.

#### AMP-T2D-GENES exome sequence genotype processing

Genotyping and quality control (QC) for AMP-T2D-GENES samples were performed as previously described^28^. Briefly, we obtained BAM files (containing unaligned sequence reads) from each contributing consortium (see ‘**AMP-T2D-GENES cohort and phenotype information**’ methods) and processed them in a uniform fashion using the Picard (broadinstitute.github.io/picard/), BWA^57^, and GATK^58^ software packages. This pipeline produced single nucleotide and indel variant calls within 50bp of any region targeted for capture. For each sample at each variant site, we calculated genotype dosages and hard genotype calls based on a genotype quality (GQ) threshold of ≥20. We called X chromosome dosages treating males as haploid.

We then conducted quality control to exclude samples and variants from analysis as previously described^28^. Briefly, we computed seventeen sample quality control metrics (see Flannick *et al*., 2019 Supplementary information^28^) and excluded samples who were outliers according to one or more metrics. We then excluded variants with a low call rate (<0.3), high heterozygosity (1), or high heterozygote allele balance (100% or 0% of heterozygous genotype reads called non-reference). Following quality control, we used a set of variants with MAF >5% across each of the five ancestries to calculate genome-wide identity-by-descent (IBD) between all sample pairs, conduct a transethnic principal component analysis (PCA), and compute genetically determined sex. To produce an analysis set of unrelated, homogenous samples, we then removed additional samples according to three metrics: (a) for sample pairs inferred to be twins or duplicates across studies (*i.e.* with IBD values >0.9), we excluded the sample with the lower mean GQ value across variants; (b) we excluded samples that did not cluster within one of the five major ancestries based on visual inspection of the first two PCs; and (c) we excluded samples whose inferred genetic sex did not match their reported sex. As a final step, we then removed any variants that became monomorphic due to the sample exclusions. The resulting analysis set consisted of 45,231 individuals and 6.33M variants.

#### AMP-T2D-GENES single variant association analysis

We conducted single variant association analysis following a previously described procedure^28^. Briefly, we first stratified samples into subgroups according to cohort and (b) ancestry and/or sequencing technology within cohort (where applicable). As previously described^28^, we did not fully stratify three cohorts: (a) we stratified samples from the ESP consortium^21^ by ancestry only, (b) we stratified samples from the SIGMA consortium^53^ by sequencing technology only, (c) and we did not stratify samples from the GoT2D consortium^45^. Following stratification, there were up to 23 distinct sample subgroups with measurements for each phenotype.

We conducted single-variant association analysis for each phenotype within each subgroup using the efficient mixed-model association expedited (EMMAX) method^59^ implemented in the Efficient and Parallelizable Association Container Toolbox (EPACTS) software package (genome.sph.umich.edu/wiki/EPACTS). We computed a genetic relationship matrix for each ancestry using the set of variants with MAF >0.05 in the ancestry^28^. We included sequencing technology as a covariate for ESP samples (as ESP was stratified for ancestry only) but, as previously noted^28^, we did not include covariates for age or sex. For single-variant analysis, we collapsed multi-allelic variants into a single alternate allele.

To assess test calibration and outlier results, we inspected results for each phenotype and subgroup (N=394) visually via QQ plots and quantitatively via genomic inflation factors (*i.e.* lambda values). We found results to be well-calibrated following filtering of variants according to seven previously described^28^ metrics:

(1) Hardy-Weinberg equilibrium *p*-value (*p*(HWE)) ≤1×10^-6^
(2) call rate (CR) <0.5 and alternative allele (Alt) GQ score <90
(3) CR <0.8 and Alt GQ score <80
(4) CR <0.9 and Alt GQ score <70
(5) CR <0.9 and p(miss) <0.01
(6) multiallelic variant and p(HWE) <0.01
(7) multiallelic variant and p(miss) <0.01

Following subgroup-level single-variant association analysis, we combined results using a fixed-effect inverse-variance weighted meta-analysis implemented in the METAL software package^60^. All coding variant associations with *p*≤4.3×10^-7^ (*i.e.* significance threshold not adjusted for multiple hypothesis testing) and non-coding variant associations with *p*≤5×10^-8^ (*i.e.* also not adjusted for multiple hypothesis testing) for any phenotype are listed in **Supplementary Table 13**.

For variants on the X chromosome, we analyzed males and females separately within each subgroup, assuming males as haploid and females as diploid. We then combined results across male and females using a fixed-effect inverse-variance weighted meta-analysis (implemented via METAL^60^) before conducting the subgroup-level meta-analyses.

#### AMP-T2D-GENES gene-level association analysis

As for single-variant association analysis, we conducted gene-level association analysis following a previously described procedure^28^. For each gene, we used the EPACTS software package to perform burden tests across seven variant masks (see ‘**Gene-level grouping**’ methods). We ran burden tests on a set of unrelated samples (all pairs IBD <0.25) with 10 transethnic PCs, sample subgroup (see ‘**AMP-T2D-GENES single variant association analysis**’ methods), and sequencing technology as covariates. For reasons previously described^28^, we performed the burden testing as a single “mega-analysis” across all samples and not a meta-analysis of sample subgroups (as done for single variant association analysis). Nonetheless, we still performed variant filtering at the sample subgroup level by setting genotypes for an entire subgroup as missing during association testing. Unlike single-variant association analysis, for which we collapsed alternate alleles for multi-allelic variants, for gene-level testing we included in each mask only the allele(s) that passed the filters defined for the mask. For genes on the X chromosome, we analyzed males and females separately before combining results via a fixed-effect inverse-variance weighted meta-analysis (implemented via METAL^60^). All genes with *p*≤2.5×10^-6^ associations (*i.e.* threshold not adjusted for multiple hypothesis testing) for any phenotype and mask are listed in **Supplementary Table 14**.

#### AMP-T2D-GENES SNP array analysis

To construct common variant PS values for AMP-T2D-GENES samples, we used SNP array data generated by the GoT2D^45^, SIGMA^53^, and T2D-GENES^28, 45^ consortia. As previously described^28^, we aggregated available SNP array data and then conducted genotype imputation with the Michigan Imputation Server^61^. Genotypes with CR >95%, *p*(HWE)>1×10^-6^, differential genotype CR between cases and controls *p* >10^-5^, and with MAF >1% were included in autosomal variant imputation. As previously noted^28^, we used two reference panels for imputation: (a) the Haplotype Reference Consortium Panel^62^ for Europeans (N=32,470 individuals) and (b) the 1000 Genomes Phase 3 Panel^63^ for non-Europeans (1000G; N=2,504 individuals). Following imputation, we applied the same set of sample and variant quality control metrics as we used for exome sequence analysis (see ‘**AMP-T2D-GENES exome sequencing genotype processing**’ methods).

### UK Biobank Analysis

#### UK Biobank phenotype information

The UK Biobank (UKB) is a prospective cohort of individuals from the general population of the United Kingdom with ages that range from 40 to 69 years. It contains extensive and publicly available genotype, phenotype, and healthcare record data^27^. We analyzed 10 quantitative phenotypes from the UKB: BMI, Diastolic-BP, Systolic-BP, Total-C, triglyceride, LDL-C, HDL-C, Waist:Hip, HbA1C, and red blood cell count (RBC). For HbA1C, only T2D-free control subjects were included in the analysis (see ‘**UK Biobank type 2 diabetes case definition**’ below). As for AMP-T2D-GENES, sample sizes varied depending on the phenotype (**Supplementary Table 1**). Statistics for each phenotype are presented in **Supplementary Table 15**.

#### UK Biobank phenotype transformations

Prior to association analysis, we first normalized phenotypes using winsorization to remove extreme values falling outside of 5 standard deviations from the phenotype distribution mean. We then used linear regression to adjust each phenotype for age, age^2^, and sex, performed inverse-normalization of the calculated residuals, and we finally scaled phenotype values to the original measurement units by multiplying them by the standard deviation of the original phenotype distribution^56^.

#### UK Biobank type 2 diabetes case definition

We defined T2D case status in the UKB using a previously published algorithm based on diabetes diagnosis, diabetes medication use, and age at diagnosis^65^. As previously noted^64^, the case definition also included touchscreen self-reported diagnosis as well as diagnosis and medication information provided at repeat visits. To reduce the potential that cases in fact had type 1 diabetes, we removed individuals from analysis if their reported “age of diabetes diagnosis” was ≤ 40 years.

#### UK Biobank genotype processing

We analyzed UKB exome sequence data from the first release of ∼50,000 samples^26^ – specifically, the Plink binary file set produced by the functionally equivalent (FE) pipeline. The data was first harmonized via two steps:

(1) Variants were strand-aligned to 1000 Genomes Phase 3 Version 5 variants via the Genotype Harmonizer software package^66^.
(2) Following the Genotype Harmonizer step, non-1000 Genomes variants were manually reconciled with the human reference assembly GRCh37. This step removes (a) variants that do not match a reference allele and (b) monomorphic variants (all individuals carrying the reference allele). Multiallelic variants were retained if an allele matched the reference.

Following harmonization, we removed variants with CR <0.3 or heterozygosity=1 across all samples.

#### UK Biobank Ancestry Inference

We extracted 5,583 variants observed in both the UKB dataset and the 1,000 Genomes (phase 3, version 5) reference panel. We used these variants to calculate PCs of a combined UKB and 1,000 Genomes dataset via the FlashPCA2 software^67^. We then used the software Klustakwik (http://klustakwik.sourceforge.net/) to perform unsupervised classification on the first 3 PCs. This software uses Gaussian mixture

modeling to identify normally distributed clusters of samples. We then labeled these clusters according to the (known) ancestries of the 1,000 Genomes samples within them (African, Admixed American, East Asian, European, or South Asian). We removed UKB samples (N=5) that did not fit into any of the clusters from further analysis (**Supplementary Table 16**).

#### UK Biobank sample-level quality control

Following the sample QC procedure we developed for the AMP-T2D-GENES study, we evaluated 10 metrics for each UKB sample using the Hail (hail.is) software package:

(1) number of heterozygous variants + number of homozygous variants (n_non_ref)
(2) number of heterozygous variants (n_het)
(3) number of homozygous reference alleles + number of heterozygous variants + number of homozygous variants (n_called)
(4) fraction of variants with called genotypes (call_rate)
(5) transition/transversion ratio (r_ti_tv)
(6) inbreeding coefficient (het)
(7) inbreeding coefficient for variants with MAF ≥
(8) inbreeding coefficient for variants with MAF <0.03 (het_low)
(9) number of homozygous alternate variants (n_hom_var)
(10) heterozygous / homozygous variant ratio across all variants (r_het_hom_var)

To account for the effects of population substructure on these metrics, we adjusted each metric according to the top 3 genetic PCs. To identify outlier samples, we clustered each sample metric into Gaussian distributed subsets using the Klustakwik software package (http://klustakwik.sourceforge.net/); we then removed outlier samples that did not fit into one of the Gaussian distributed subsets. For the metrics n_called and call_rate, we removed only outlier samples below the mean of the distribution.

In addition to excluding samples who were outliers according to individual sample metrics, we also excluded samples that were outliers according to combinations of sample metrics. To identify these outliers, we first calculated PCs across the metrics that explained 95% of their variation (we did not include the ’call_rate’ and ’n_called’ metrics in this analysis as they were characterized by long distributional tails inconsistent with the other metrics). We then scanned for samples that exhibited deviation from any of the metric PCs: as we did for the individual metrics, we used Klustakwik (http://klustakwik.sourceforge.net/) to identify samples that lay outside of the distribution of one of more metric PCs. In total, we removed 76 samples by this analysis (**Supplementary Table 16**).

We also assessed potential discordance between each sample’s reported sex and its genetically determined sex using the HAIL software package (hail.is). We did not remove any additional samples (N=0) due to discordance between reported and genetic sex, although we did remove N=16 samples whose imputation for sex failed (*i.e.* flagged as a ‘PROBLEM’ by Hail)(**Supplementary Table 16**).

#### UK Biobank variant-level quality control

We assessed variant quality using call rate and HWE calculated within EUR samples. We excluded variants with HWE *p*<1x10^-6^ or call rate <0.3 from further analysis. In total, 62,420 variants were flagged for removal resulting in a total of 8.89M variants remaining for downstream analysis.

#### UK Biobank kinship analysis

To produce a set of variants for inferring genetic relationships among samples, we first used the Plink software package^68^ to remove non-autosomal (--chr flag), non-A/C/G/T (--snps-only flag), low call rate (CR ≤ frequency (--maf flag) variants. We also removed variants with positions in known high LD regions (http://genome.sph.umich.edu/wiki/Regions_of_high_linkage_disequilibrium_(LD)). We then LD-pruned the remaining variants with Plink to produce a set of 57,297 variants.

We used the KING relationship inference software package^69^ to determine sample pair kinship coefficients. We considered samples with kinship coefficients >0.4 as potential duplicates. If the clinical data for the duplicate pair were identical, then we removed the sample with the lower genotype call rate from analysis. If the clinical data were not identical, we removed both samples. In addition to removing duplicate samples, we also checked for any individuals that exhibited kinship values indicating a 2nd degree relative or higher relationship with 10 or more others and found none to be removed (N=0).

#### UK Biobank single variant association analysis

Single variant association analysis in the UKB was performed using the HAIL linear regression function (*i.e.* the “hail.methods.linear_regression_rows()” function) to assess association between each variant and each inverse-normal transformed phenotype. The single variant regression model included covariates for the first nine PCs of ancestry but not age and sex (as these were included when transforming the phenotypes). The analysis included only variants with ≥3 minor allele counts. We analyzed variants on the X-chromosome analogously to those in AMP-T2D-GENES. All coding variants with *p*≤4.3×10^-7^ and non-coding variants with *p*≤5×10^-8^ for any phenotype are listed in **Supplementary Table 17**.

#### UK Biobank gene-level association analysis

As for the AMP-T2D-GENES study (see ‘**AMP-T2D-GENES gene-level association analysis**’ methods), we performed gene-level association analysis using the EPACTS software package. For each gene, we conducted fixed-effect burden tests for seven variant masks (see ‘**Gene-level grouping**’ methods). Burden test models were adjusted for the first 9 ancestry-specific PCs as model covariates. Genes on the X-chromosome were analyzed analogously to those in AMP-T2D-GENES. All gene-level associations with *p*≤2.5×10^-6^ for any phenotype and mask are listed in **Supplementary Table 18.**

### Variant annotation

We used the variant effect predictor (VEP) software package^70, 71^ to produce variant annotations for the AMP-T2D-GENES and UKB datasets. We ran VEP with two plugins: (a) the LofTee plugin^72^, which predicts loss-of-function variants, and (b) the dbNSFP plugin (version 3.2)^73^, which produces annotations from 15 bioinformatic algorithms. Variant annotations were produced across all ENSEMBL transcripts via the “--flag-pick-allele” option^74^ using a previously described default “ordering criteria” for transcripts^28^. For annotating results from the single variant analysis, we used the “best guess” annotation based on the first transcript according to the ordering criteria.

### Gene-level grouping

We used the variant annotations and previously published criteria^28^ to group variants found in the AMP-T2D-GENES and UKB studies into seven different groupings, termed “variant masks.” The seven masks were ordered (with 1 being most strict) by the likelihood of the deleteriousness of variants within them:

(1) “LoFTee” consists of variants predicted to be loss of function with high confidence according to the LOFTEE plugin
(2) “16/16” includes variants in the previous mask (LoFTee) and variants with VEST3_rankscore ≥ 0.9 and CADD_raw_rankscore ≥ 0.9 and DANN_rankscore ≥ 0.9 and Eigen_PC_raw_rankscore ≥ 0.9 and FATHMM_pred=D and FATHMM_MKL_pred=D and PROVEAN_pred=D and MetaSVM_pred=D and MetaLR_pred=D and M_CAP_score ≥ 0.025 and Polyphen2_HDIV_pred=D and Polyphen2_HVAR_pred=D and SIFT_pred=D and LRT_pred=D and MutationTaster_pred=D or A
(3) “11/11” includes variants in the previous mask (16/16) and variants with FATHMM_pred=D and FATHMM_MKL_pred=D and PROVEAN_pred=D and MetaSVM_pred=D and MetaLR_pred=D and M_CAP_score ≥ 0.025 and Polyphen2_HDIV_pred=D and Polyphen2_HVAR_pred=D and SIFT_pred=D and LRT_pred=D and MutationTaster_pred=D or A
(4) “5/5” includes variants in the previous mask (11/11 ) and variants with Polyphen2_HDIV_pred=D and Polyphen2_HVAR_pred=D and SIFT_pred=D and LRT_pred=D and MutationTaster_pred=D or A
(5) “5/5 LoFTee LC 1%” includes variants in the previous mask (5/5) and MAF<1% variants predicted to be loss of function with low confidence according to the LOFTEE plugin
(6) “1/5 1%” includes variants in the previous mask (5/5 LoFTee LC 1%) and MAF<1% variants with either Polyphen2_HDIV_pred=D or Polyphen2_HVAR_pred=D or SIFT_pred=D or LRT_pred=D or MutationTaster_pred=D or A
(7) “0/5 1%” includes variants in the previous mask (1/5 1%) and all missense variants with MAF < 1%.

### Single variant meta-analysis of AMP-T2D-GENES and the UKB

We combined single variant association results across AMP-T2D-GENES and UKB via a sample size weighted meta-analysis using the METAL software package^60^. We chose a sample size weighted meta-analysis instead of an inverse variance weighted meta-analysis to avoid any issues with differing phenotype units between the two studies. For coding variants, we used a significance threshold of *p*≤1.8×10^-8^ (*p*≤4.3×10^-7^ with Bonferroni correction for 24 phenotypes^75^); for noncoding variants, we used a significance threshold of *p*≤2.1×10^-9^ (*p*≤5×10^-8^ with Bonferroni correction for 24 phenotypes). All variants with significant associations across all phenotypes are listed in Supplementary Table 2.

### Gene-level meta-analysis of AMP-T2D-GENES and the UKB

We meta-analyzed the gene-level association results from AMP-T2D-GENES and UKB for each of the seven aforementioned masks by conducting sample-size weighted meta-analyses using the METAL^60^ software package. These meta-analyses produced up to seven *p*-values per gene (*i.e.* one *p*-value for each mask following meta-analysis); mask-level meta-analysis results are presented in **Supplementary Table 19**. We then used a previously published strategy^28^ to consolidate *p*-values across masks by (a) assigning the gene the lowest *p*-value across masks; (b) calculating the effective number of gene-level tests for the gene according to the independence of the variants across its masks; and (c) correcting the *p*-value for the effective number of tests. QQ plots and genomic inflation factors (λ) of the consolidated *p*-values showed that the tests were, if anything, conservative (all λ <1).

For the gene-level meta-analysis, we considered genes with *p*≤1.0×10^-7^ (*p*≤2.5×10^-6^ with Bonferroni correction for 24 phenotypes) to be exome-wide significant. All gene-level associations that reach this threshold are listed in **Supplementary Table 4**.

### Calculating genetic effect sizes for raw HbA1C

To calculate association effect sizes in clinically-relevant units, as done in past studies^6^, we recomputed single-variant and gene-level HbA1C associations using raw HbA1C levels (*i.e.* percent glycated hemoglobin) as opposed to inverse-normalized values. Analysis protocols were identical except for the HbA1C values used. As HbA1C units reported in the UKB dataset are in ‘mmol/mol’, we converted these to ‘percent glycated hemoglobin’ units using

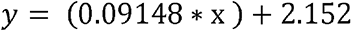

where *y* is the HbA1C reported as percent glycated hemoglobin and *x* is HbA1C reported in mmol/mol.

Single variant association results using raw HbA1C as the outcome variable are listed in **Supplementary Table 3**, while gene-level association results raw HbA1C as the outcome variable are presented in **Supplementary Table 5**. In the manuscript text, all gene-level effect sizes reported correspond to the variant mask with the smallest *p*-value for the gene.

### Proportion of variance explained calculations

The proportion of variance explained (PVE) either by individual SNPs or by a collection of variants within a gene were calculated using genetic association summary statistics and following a previously derived formula^76^:

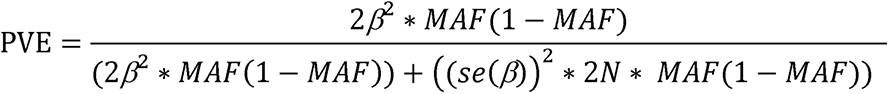

where, for single variants, β is the effect size of the genetic association, se(β) is the standard error of β, MAF is minor allele frequency, and N is sample size. For MAF values for gene-level associations, we used the aggregate minor allele frequency across all rare variants included in the associations. For variants and/or genes found on the X chromosome, we treated males as haploid.

As we found the *PIEZO1* rare variant gene-level association to explain a larger proportion of variance than a previously identified nearby common variant association (rs837763)^6^, we evaluated whether LD between the common variant and rare variants included in the gene-level analysis impacted the estimated variance explained. To do so, we re-conducted the gene-level analysis for the mask that produced the smallest *p*-value (5/5 mask; see ‘**Gene-level grouping**’ methods) conditional on the common variant rs837763. For AMP-T2D-GENES, we included only individuals with SNP array data (N=16,701) in this analysis. We then evaluated, within each ancestry, the proportion of variance explained by *PIEZO1* with and without inclusion of rs837763 as a covariate. Results suggested that including rs837763 as a covariate had little impact on the estimated proportion of variance explained by the *PIEZO1* rare variant gene-level association (**Supplementary Table 6**).

### Defining GWAS loci for rare variant association analysis

To assess whether variants or genes identified during rare variant association analysis were located within known GWAS loci, we calculated the genomic distance between a variant (or gene) and the nearest genome-wide significant association (*p*≤5×10^-8^) for the phenotype of interest. We extracted genome-wide significant associations for each phenotype from the Type 2 Diabetes Knowledge Portal^77^. We considered variants or genes within 125kb of a genome-wide significant association to be within a known GWAS locus.

### Gene set enrichment analyses

We conducted enrichment analyses following a previously described procedure^28^. Specifically, to test enrichment for a given set of genes, we (a) calculated the exome-wide percentile for each gene according to its gene-level association *p*-value (with lower *p*-values translating to higher percentiles); (b) matched each gene to a set of background genes with similar properties (*e.g.* number of variants, combined allele count); and (c) conducted a one-sided Wilcoxon rank sum test to test whether genes within the gene set had higher percentiles (*i.e.* lower *p*-values) than expected by chance. We drew gene-level association *p*-values from the gene-level meta-analysis results (see ‘**Gene-level meta-analysis of AMP-T2D-GENES and the UKB**’ methods), following application of the minimum *p*-value consolidation method.

We conducted enrichment tests for two types of gene sets. First, to test whether genes with a pre-specified annotation had stronger than expected associations, we curated pre-defined gene sets based on knockout mouse phenotypes (see ‘**Mouse gene set definitions**’ methods). Second, to test whether genes associated with HbA1C were likewise associated with RBC, or vice versa, we constructed gene sets from the top (meaning smallest *p*-value) *n* HbA1C (or RBC) gene-level associations, with *n* ranging from 1 to 1,000. For each *n*, we then tested for enrichment either for (a) stronger RBC associations (among the top HbA1C-associated genes) or (b) stronger HbA1C associations (among the top RBC-associated genes). We used the Benjamini and Hochberg method^78^ to correct the enrichment *p*-values for multiple hypothesis testing.

### Mouse gene set definitions

To identify erythrocytic gene sets based on knockout mouse annotations, we searched the mouse genome informatics (MGI) database for genes annotated with terms including ‘erythrocyte’, ‘erythropoiesis’, or ‘hematocrit’. The resulting erythrocytic gene sets (N=10) include: 1) abnormal erythrocyte cell number (N=10), 2) abnormal erythrocyte morphology (N=105), 3) abnormal erythrocyte osmotic lysis (N=36), 4) abnormal erythrocyte physiology (N=33), 5) abnormal erythropoiesis (N=155), 6) decreased erythrocyte cell number (N=237), 7) decreased hematocrit (N=205), 8) increased erythrocyte cell number (N=85), 8) increased hematocrit (N=89), 9) increased nucleated erythrocyte cell number (N=47), 10) low mean erythrocyte cell number (N=16). To identify glycemic gene sets, we searched for genes annotated with terms including ‘glucose’, ‘insulin’, or ‘diabetes.’ The resulting glycemic gene sets (N=25) include 1) abnormal circulating glucose level (N=43), 2) abnormal circulating insulin level (N=30), 3) abnormal glucose homeostasis (N=245), 4) abnormal glucose tolerance (N=25), 5) abnormal insulin secretion (N=22), 6) decreased adipocyte glucose uptake (N=21), 7) decreased circulating glucose level (N=385), 8) decreased circulating insulin-like growth factor I level (N=61), 9) decreased circulating insulin level (N=360), 10) decreased fasting circulating glucose level (N=21), 11) decreased insulin secretion (N=139), 12) decreased muscle cell glucose uptake (N=27), 13) impaired glucose tolerance (N=363), 14) improved glucose tolerance (N=284), 15) increased adipocyte glucose uptake (N=26), 16) increased cardiac cell glucose uptake (N=13), 17) increased circulating glucose level (N=223), 18) increased circulating insulin-like growth factor 1 level (N=13), 19) increased circulating insulin level (N=228), 20) increased fasting circulating glucose level (N=116), 21) increased insulin secretion (N=53), 22) increased insulin sensitivity (N=197), 23) increased muscle cell glucose uptake (N=36), 24) increased urine glucose level (N=30), 25) insulin resistance (N=201).

### Directional concordance of genes within enriched gene sets

To test whether the direction of effect on HbA1C was consistent across gene-level associations within a gene set, we first filtered to genes with HbA1C association (*p*≤0.05). We then assigned a direction of effect to each gene according to the sign of the effect size from the most significant (*i.e.* smallest *p*-value) mask.

We then tested for directional concordance among these filtered genes by evaluating, via a binomial test, whether there were more negative effect sizes than the 50% expected by chance. We conducted this analysis for significantly enriched erythrocytic and glycemic gene sets. We also evaluated concordance both (a) without removing overlapping genes found in multiple gene sets and (b) after removing overlapping genes found in multiple gene sets. In both cases, we considered *p*≤0.05 as significant.

### Polygenic adjustment scores for HbA1C

We calculated polygenic scores (PS) similarly for both common and rare variants. For the common variant PS, we used a set of variants and effect sizes from a previous GWAS publication^6^. The PS was calculated as

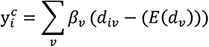

where 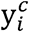is the common variant adjustment for individual *i,βv* is the estimated effect derived for a particular variant (*v*) included in the PS, *d_iν_* is the allelic dosage for variant *v* for individual *i*, and *E(d_ν_*) is the minor allele frequency of *v*.To obtain the adjusted HbA1C value for individual *i*, we subtract the calculated PS value 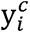. from the individual’s reported HbA1C.

For the rare variant PS, the key distinction relative to the common variant PS is the set of variants over which the summation above is taken. In contrast to the common variant PS, for which the set of variants included is fixed across individuals, the set of variants included in the rare variant PS can vary across individuals and is potentially unbounded – the summation is (in principle) taken over all rare variants that could ever be observed in a population. We resolve this issue by assigning rare variants to a set of pre-defined groups *g*, for each of which the needed parameters are constant and estimable at the time the model is built. The rare variant PS then becomes

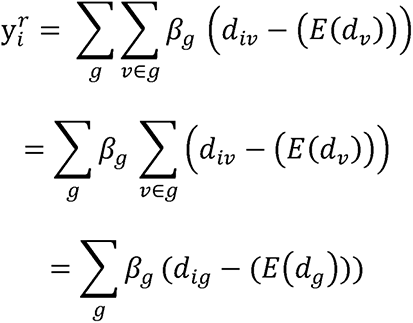

where β*_g_* is the estimated effect derived for variants within group *g*, *d_ig_* is the combined allelic dosage for all variants in the group, and *E*(*d_g_*) is the combined minor allele frequency of variants in *g*. As for the common variant PS, to obtain the adjusted HbA1C value for individual *i*, we subtract the calculated PS value 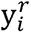. from the individual’s reported HbA1C.

### Genes included in the rare variant polygenic score

To determine genes to be included in the rare variant PS, we filtered genes according to (a) their likelihood of harboring a true association with HbA1C and (b) their likelihood of impacting HbA1C through erythrocytic pathways. We considered genes to have evidence of impacting HbA1C through erythrocytic pathways if they satisfied at least one of three criteria: they either (a) were included in an erythrocytic mouse gene set that showed nominally significant (*p*<0.05) enrichment for HbA1C associations (see ‘**Gene set enrichment analyses**’ methods); (b) demonstrated a rare variant gene-level association at nominal significance (*p*≤0.05) with RBC; or (c) were located within an RBC GWAS locus (see ‘**Defining GWAS loci for rare variant association analysis**’ methods).

In total, we explored five strategies for determining genes (**Figure 3a**):

(1) *Strict gene set*. These genes demonstrated evidence of impacting HbA1C through erythrocytic pathways (as defined above) and, as evidence of HbA1C association, demonstrated exome-wide significant gene-level rare variant HbA1C associations.
(2) *Relaxed gene set*. These genes demonstrated evidence of impacting HbA1C through erythrocytic pathways (as defined above) and, as evidence of HbA1C association, demonstrated nominally-significant (*p*≤0.05) HbA1C gene-level rare variant associations and at least one further line of evidence supporting their association with HbA1C: (a) presence in a glycemic mouse gene set with *p*<0.05 HbA1C enrichment (see ‘**Gene set enrichment analyses**’) or (b) proximity to an HbA1C GWAS association (see ‘**Defining GWAS loci for rare variant association analysis**’ methods).
(3) *Loose gene set.* These genes demonstrated evidence of impacting HbA1C through erythrocytic pathways (as defined above) and, as evidence of HbA1C association, demonstrated nominally-significant (*p*≤0.05) HbA1C gene-level rare variant associations.
(4) *Nominal gene set*. These genes demonstrated nominally-significant (*p*≤0.05) HbA1C rare variant gene-level associations. They were not required to demonstrate evidence of impacting HbA1C through erythrocytic pathways.
(5) *Negative control gene set*. These genes demonstrated nominally-significant (*p*≤0.05) HbA1C gene-level rare variant associations and were (a) included in a glycemic gene set that showed nominally significant (*p*<0.05) enrichment for HbA1C associations (see ‘**Gene set enrichment analyses**’ methods) or (b) were located within a fasting glucose or fasting insulin GWAS locus (see ‘**Defining GWAS loci for rare variant association analysis**’ methods). Genes with evidence of impacting HbA1C through erythrocytic pathways (*i.e.* found ‘strict’, ‘relaxed’, or ‘loose’ gene set) were omitted.

### Variant weights in the rare variant polygenic score

After we define a set of genes for the rare variant PS, we then define a set of variant groups *g*(*e*) for each gene *e*, estimated effect sizes *β_g_*(e) for each group *g*(*e*), and the estimated combined allele frequency in the population of variants in *g*(*e*). We tested three different methods for determining these parameters: 1) the nested variant method, 2) the unique variant method, and 3) the weighted variant method (**Figure 3b**). In all cases, gene-level effect sizes determined using these methods were estimated within the UKB, our discovery sample.

#### Nested variant method

Intuitively, the nested method assigns an effect to *v* equal to the aggregate effect size for the most stringent mask including *v*. The seven masks we analyzed (see ‘**Gene-level grouping**’ methods) are ordered in decreasing stringency as LofTee, 16/16, 11/11, 5/5, 5/5 LofTee LC 1%, 1/5 1%, 0/5 1%. Masks are nested in that more stringent masks are subsets of less stringent masks; therefore if a variant is included in a mask it is also included in all less stringent masks. The nested method uses one group *g*(*e*) for each gene and each mask, each consisting of the variants within that gene and mask but not within any more stringent masks. The combined minor allele frequency *E*(*d_g_*_(*e*)_) is equal to the fraction of individuals that carry variants in *g*(*e*). The effect size *β_g_*_(*e*)_ is equal to the aggregate effect size across all variants in the gene and mask – importantly, this includes variants also in more stringent masks and therefore not in *g*(*e*).

#### Unique Variant Method

The unique variant method is similar to the nested variant method but attempts to correct for the potential overestimation of effects resulting from variants within more stringent masks. The unique variant method uses the same groups as the nested variant method: one group *g*(*e*) for each gene and mask, each consisting of the variants within that mask but not within any more stringent masks. The combined minor allele frequency *E*(*d_g_*_(*e*)_) is equal to the fraction of individuals that carry variants in *g*(*e*). The effect size *β_g_*_(*e*)_ is equal to the aggregate effect size across all variants in the gene unique to the mask – in contrast to the nested method, this includes only variants in *g*(*e*) and therefore excludes variants also in more stringent masks.

#### Weighted Variant Method

The nested and unique methods each have potential limitations: the nested method may overestimate variant effect sizes by including all variants in a mask (including those in more stringent masks), while the unique method may produce noisy effect size estimates by reducing the number of variants included in effect size estimation. The weighted variant method attempts to estimate a maximum effect size for a gene using data from all masks (corresponding to the effect size for complete loss of function) and to then derive group-specific weights by multiplying this overall effect size by the estimated proportion of loss-of-function variants in the group. Following a previous study^28^, we assigned each mask a weight ranging from 0 to 1, based on modeling the frequency distribution of variants in the mask as a mixture of the frequency distribution of LoFTee variants (weight 1) and the frequency distribution of synonymous variants (weight 0). We then, for each gene, conducted a weighted burden test in which HbA1C was regressed on all coding variants, each weighted by the value of their most stringent mask. The resulting “weighted” gene-level effect size estimate is in units of HbA1C change per loss-of-function allele. The weighted variant method then uses the same groups as the nested and unique variant method (one group *g*(*e*) for each gene and mask, each consisting of the variants within that mask but not within any more stringent masks). The combined minor allele frequency *E*(*d_g_*_(*e*)_) is equal to the fraction of individuals that carry variants in *g*(*e*). The effect size *β_g_*_(*e*)_ is equal to the weight of the mask multiplied by the weighted effect size estimate for the gene.

### Assessing accuracy of the rare variant HbA1C polygenic score

To evaluate the rare variant PS, we identified individuals who have reported HbA1C <6.5% but, after accounting for the effects of the PS, would be “reclassified” as a T2D case according to their adjusted HbA1C. To identify these individuals, we first excluded all individuals who (a) reported usage of antihyperglycemic medication or (b) did not have antihyperglycemic medication data available. The total number of individuals reclassified was taken as a measure of the sensitivity of the PS.

To measure the specificity of the PS, we then calculated, of the individuals reclassified by the PS, how many were “true” T2D cases diagnosed based on criteria not involving HbA1C or antihyperglycemic medication use – that is, either having (a) a 2-hour glucose measure ≥11.1 mmol/L or (b) fasting glucose levels ≥7 mmol.

We then calculated the ratio of reclassified true cases (*i.e.* true positives) to reclassified controls (*i.e.* false positives) and compared this to the expected ratio of true cases to controls based on the HbA1C levels of the reclassified individuals. We controlled for HbA1C within the expected ratio because individuals with HbA1C values closer to 6.5% are more likely to be cases *a priori* and, therefore, the expected ratio of reclassified cases to controls is confounded with the size of the adjustment. To calculate the expected case to control ratio, we randomly matched up to 10 individuals per reclassified individual with (a) an identical reported HbA1C level and (b) the same cohort and ancestry. We then used a Fisher’s exact test to evaluate whether the ratio of true cases to controls reclassified by the PS was significantly greater than the expected ratio. In addition to a *p*-value, the Fisher’s test produces an odds ratio (OR), which represents the fold-increase in the odds of reclassifying a true case based on the PS relative to the odds of randomly sampling a true case with the same HbA1C. We calculated ORs both within each ancestry and as well as across ancestries; trans-ethnic ORs were calculated by pooling all samples. A *p*-value ≤ significant. Reclassification results from the rare variant PS are displayed in Supplementary Table 8.

### Common variant polygenic score

We constructed the common variant PS using a previously published set of variants likely to impact HbA1C through erythrocyte pathways (**Supplementary Figure 3**)^6^. We implemented the common variant PS within the AMP-T2D-GENES study using imputed SNP array data available for 16,701 samples^28^ (See ‘**AMP-T2D-GENES SNP array analysis**’ methods). We drew effect sizes and standard errors from the study that initially defined the PS^6^; all summary statistics were downloaded from www.magicinvestigators.org. We then evaluated the common variant PS using the same approach as for the rare variant PS (see ‘**Assessing accuracy of the rare variant HbA1C polygenic score**’ methods). We calculated separate odds ratios for each ancestry-specific PS (**Supplementary Figure 3**) and then combined odds ratios across ancestries via an inverse-weighted meta-analysis^60^. Reclassification results for the common variant PS are displayed in **Supplementary Table 8**.

### Combining rare and common variant polygenic scores

To assess whether combining the rare variant PS and common variant PS via an additive model is justified, we used a conditional analysis to evaluate independence between the variants used in each PS. To do so, we repeated the gene-level analysis for each variant mask used in the rare variant rare variant PS (see ‘**Gene-level grouping**’ methods) conditional upon the variants included in the common variant PS. This analysis demonstrated that the variants included in the rare variant PS are (almost entirely) independent of the variants included in the common variant PS (**Supplementary Figure 5**).

Therefore, we constructed a combined PS assuming an additive model where we summed the separate scores for each individual. To evaluate the combined PS method, we followed the same procedure as we used to evaluate the rare and common variant PS: we used individual-level data from the AMP-T2D-GENES study to assess how many T2D cases are “reclassified” to have an adjusted HbA1C >6.5%, and whether the ratio of reclassified cases to controls is greater than the expectation from a set of individuals matched on HbA1C, cohort, and ancestry (see ‘**Assessing accuracy of the rare variant HbA1C polygenic score**’ methods).

### Polygenic score sensitivity analyses

To evaluate the extent to which the rare variant PS and common variant PS depend on individual genes and/or variants, we used a leave-one-out approach. Specifically, we repeated the PS evaluations (see ‘**Assessing accuracy of the rare variant HbA1C polygenic score**’ and ‘**Common variant polygenic score**’ methods) but with the exception that either all variants found in a gene of interest (for the rare variant PS) or a single common variant of interest (for the common variant PS) were left out of the analysis. We then compared the resulting number of reclassifications and case:control odds ratio to that of the full PS. Representative results from the leave-one-out analysis for the rare and common variant PS are displayed, respectively, in **Supplementary Table 20** and **21**.

### Scaling ancestral proportions using NHANES

As the AMP-T2D-GENES dataset was ascertained based on ancestry, its ethnic contribution is not representative of the United States (US) population. We used the most recent release (*i.e.* 2017-2018) of the National Health and Nutrition Examination Survey (NHANES) data to estimate, based on the results from the AMP-T2D-GENES dataset, how many true T2D cases in the US population would be reclassified by the various PS. First, we estimated US ancestral proportions using data from NHANES after adjusting for the survey design using the “survey” R package^79^. To then estimate the number of individuals from each ancestry in the US population who would be reclassified by the PS, we then scaled the ancestry proportions in the AMP-T2D-GENES samples to match the estimated US ancestral proportions. For this analysis, we assumed (a) the number of cases and controls would scale accordingly when adjusting for population size and (b) that the total US population size is ∼330 million based on the 2018 US Census Bureau estimates. As NHANES reports an “Asian American” sub-group, but does not further classify individuals as South or East Asian, we combined South and East Asian results from our rare and common variant PS analysis into a single “Asian” ancestry via an inverse-weighted meta-analysis via METAL^60^.

### Testing for ancestral heterogeneity

To assess heterogeneity of common or rare variant effect sizes across ancestries, we used a Cochran’s Q test as implemented in the METAL software package^60^. For the common variant PS, we tested for heterogeneity across ancestry-specific summary statistics downloaded from the original study defining the PS^6^. For the rare variant PS, we tested for heterogeneity across ancestry-specific gene-level summary statistics. To compute ancestry-specific gene-level summary statistics, we repeated our analysis procedure (see ‘**AMP-T2D-GENES gene-level association analysis**’ and ‘**UK Biobank gene-level association analysis**’ methods) and ran gene-level burden tests across each ancestry using untransformed HbA1C as the outcome across each of the seven variant masks (see ‘**Gene-level grouping**’ methods).

We assessed the resulting Q test *p*-values via QQ-plots and by calculating λ values (the median of the observed chi-square statistic from the Q test over the median of the expected chi square statistic under a null model). Larger deviations from the null model and consequent larger λ values indicate greater heterogeneity across ancestry for the tested variants or genes. The QQ plots and λ values for variants in the common variant PS and genes in the rare variant PS are displayed in **Supplementary Figure 6**.

## Supporting information

AMP-T2D-GENES Consortia Authors

## Data Availability

All data analyzed are available via (a) NCBI database of genotypes and phenotypes (dbGaP) or (b) through the UK Biobank. Genetic association summary statistics produced are available through the common metabolic diseases knowledge portal.

## Supplementary Figures

**Supplementary Figure 1.**
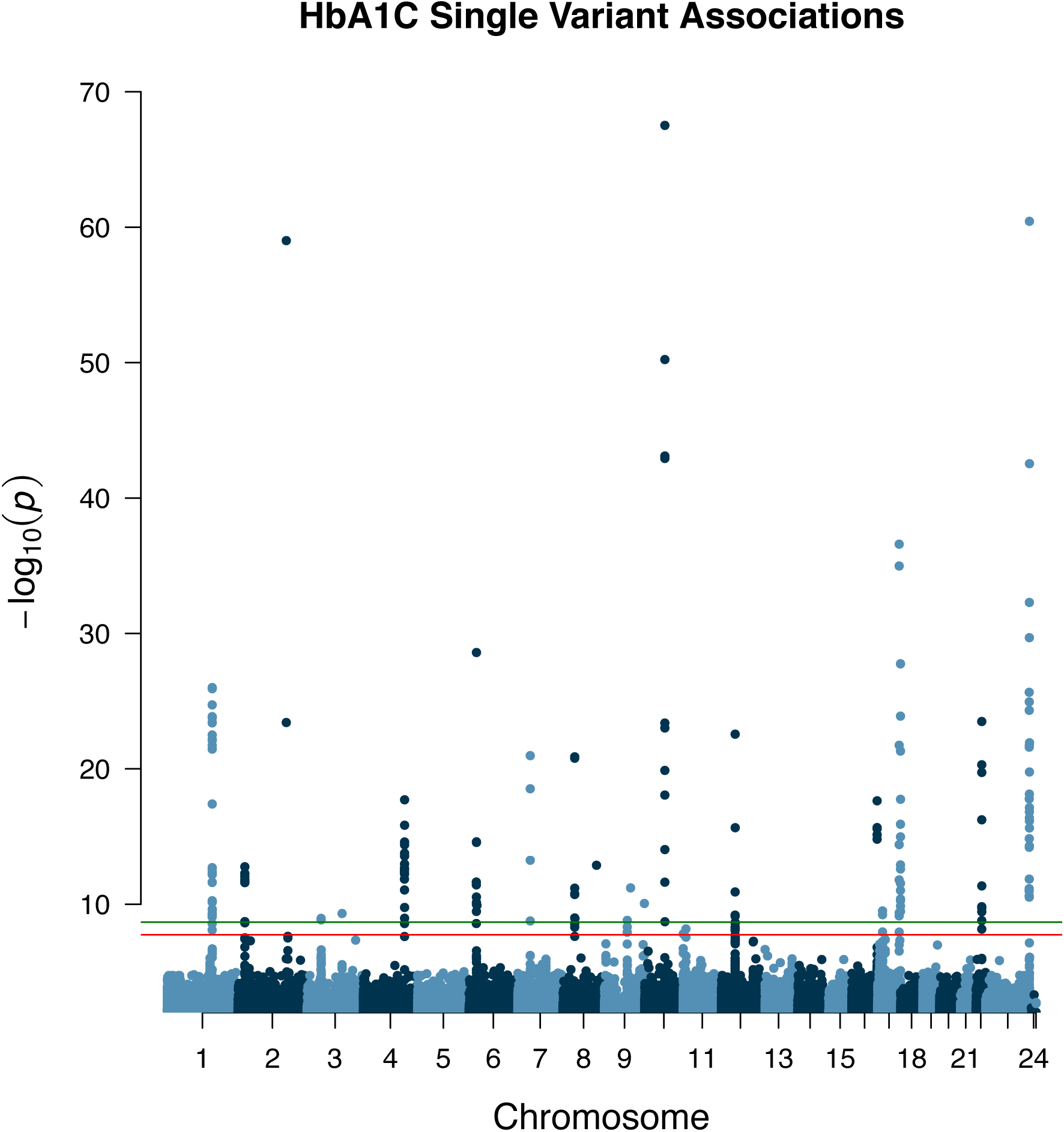
Single variant HbA1C associations. Manhattan plot of the single variant associations identified by our meta-analysis. Horizontal lines indicate the threshold used for exome-wide significance for coding variants (red; *p*≤1.8×10^-8^) and genome-wide significance for non-coding variants (green; *p*≤2.1×10^-9^).

**Supplementary Figure 2.**
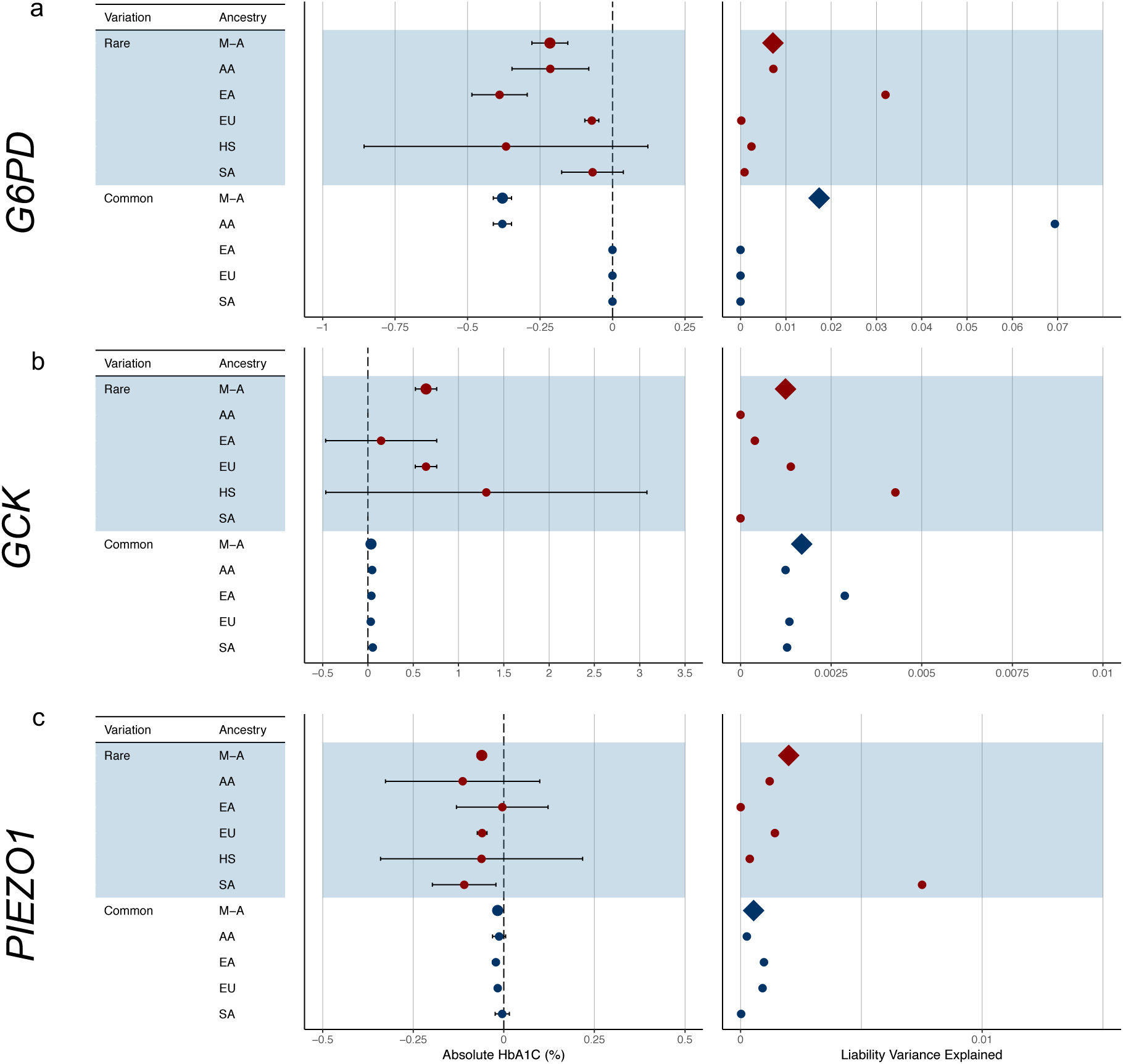
Effect sizes and proportion of variance explained for rare variant HbA1C gene-level associations. Results are displayed for **(a)** *G6PD*, **(b)** *GCK*, and **(c)** *PIEZO1*. We calculated effect sizes and liability variance explained separately for each ancestry and then combined these via a meta-analysis. We performed the calculations for the strongest associated gene-level mask and for the strongest associated common variant within 125kb of the gene. Effect sizes are displayed in percent glycated hemoglobin units; proportion of variance explained is displayed as the proportion of total liability variance (see ‘**Proportion of variance explained calculations**’ methods for details).

**Supplementary Figure 3.**
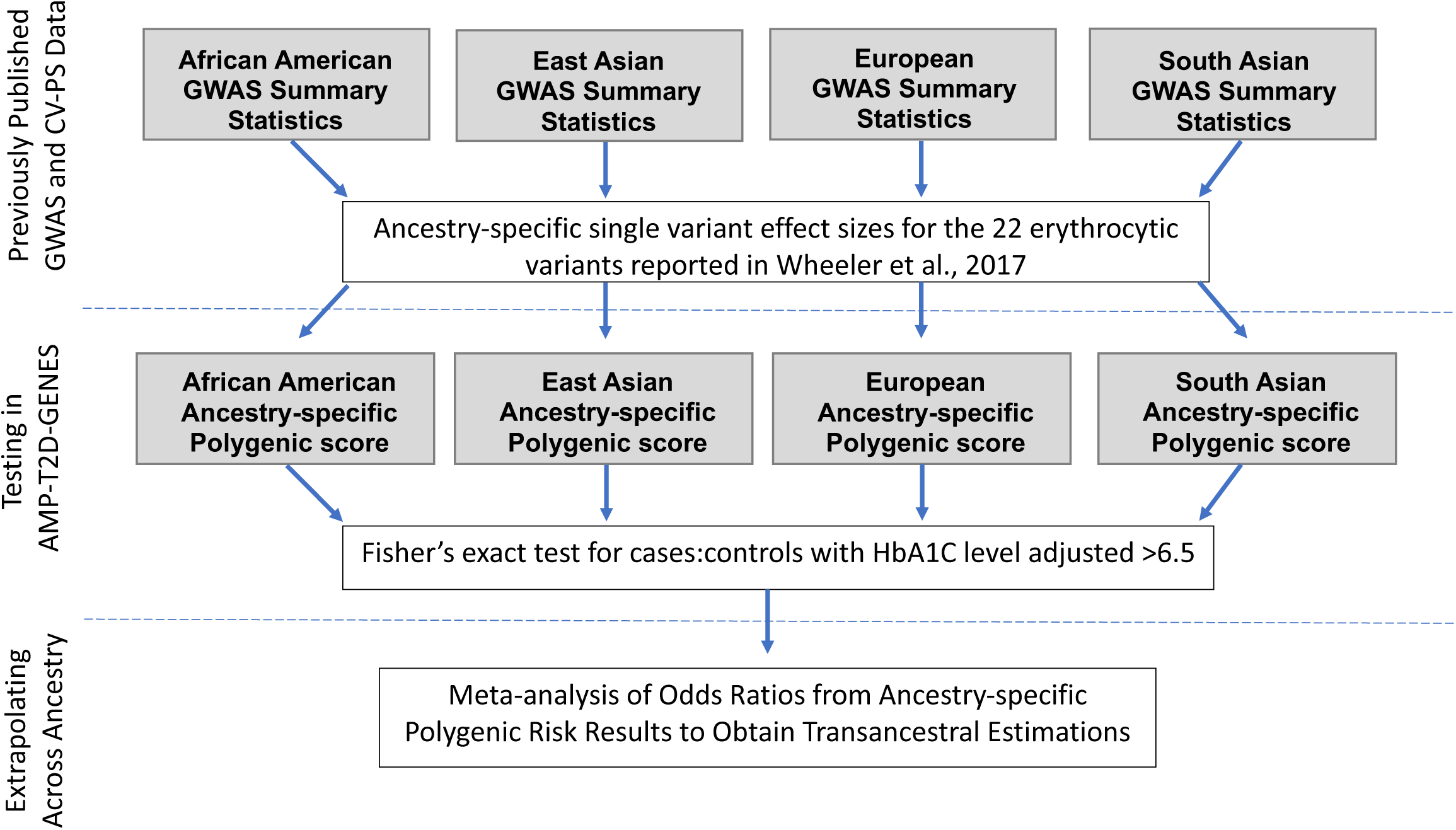
Calculating and evaluating common variant polygenic scores. We calculated common polygenic scores based on effect sizes and results from a previously published multi-ethnic HbA1C GWAS^6^. We calculated polygenic scores separately for each of the four ancestries in our test sample with available GWAS data, evaluated ancestry-specific odds ratios via a Fisher’s exact tests, and then combined these odds ratios via a fixed-effects meta-analysis to produce transethnic odds ratio (see ‘**Common variant polygenic score**’ methods for details).

**Supplementary Figure 4.**
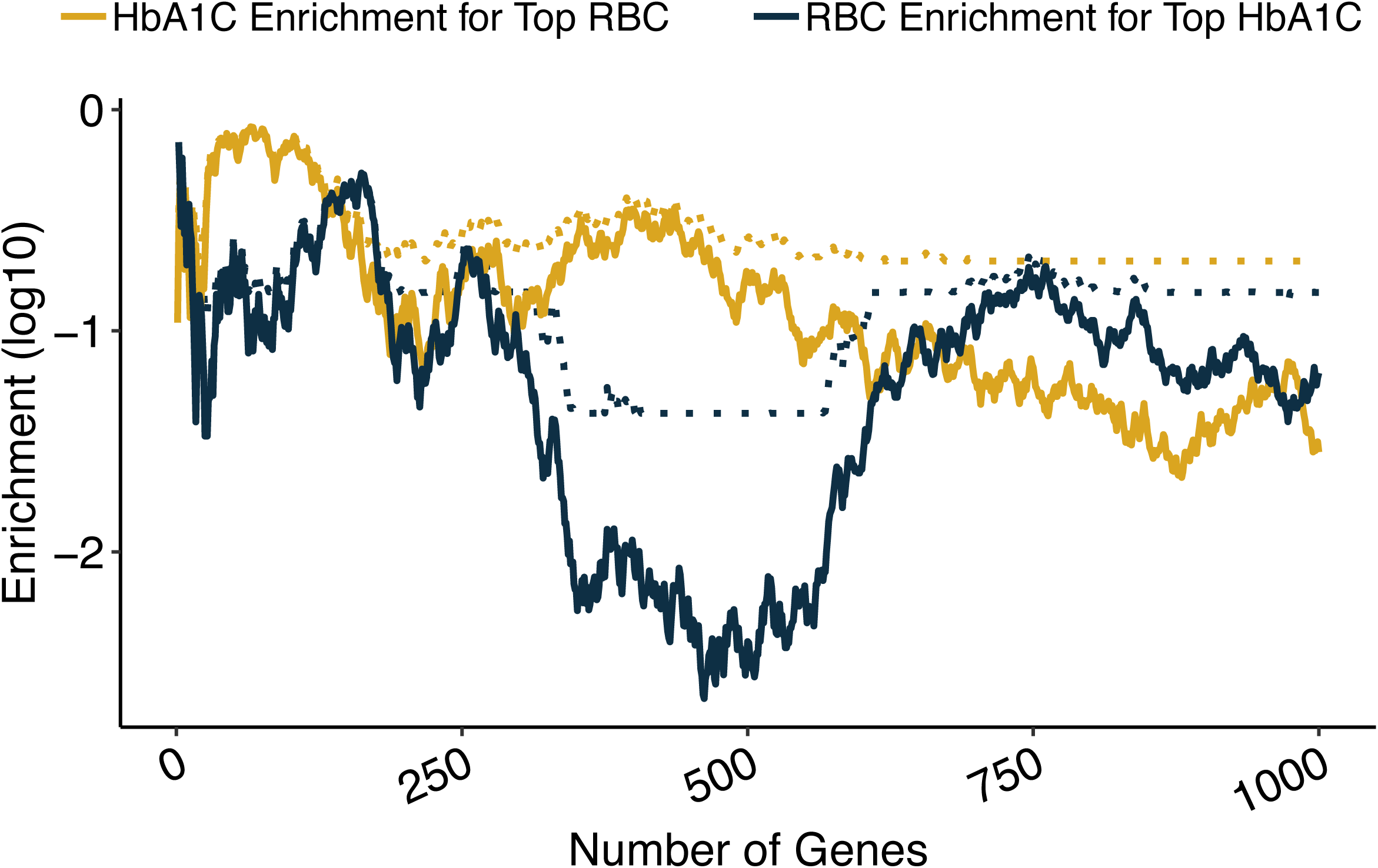
Enrichment analyses of HbA1C and RBC rare variant gene-level associations. We ranked genes by their HbA1C gene-level *p*-value and tested the degree to which the top *n* associations (with *n* ranging from 1 to 1,000) were enriched for red blood cell count (RBC) gene-level associations. Enrichments were calculated using a one-sided Wilcoxon rank-sum test, comparing the RBC gene-level *p*-values of the top *n* HbA1C associations to the RBC gene-level *p*-values of background genes matched on the number of variants and total allele count (see ‘**Gene set enrichment analyses**’ methods for details). The solid blue line in the plot shows the Wilcoxon *p*-values as a function of *n*; the dotted blue line indicates the *q*-value calculated via a Benjamini-Hochberg adjustment. As a negative control, we also conducted the reciprocal analysis in which we tested the top *i* RBC associations for enrichment for HbA1C associations; the solid yellow indicates the Wilcoxon *p*-values and dotted yellow line indicates the *q*-value as determined via the Benjamini-Hochberg method.

**Supplementary Figure 5.**
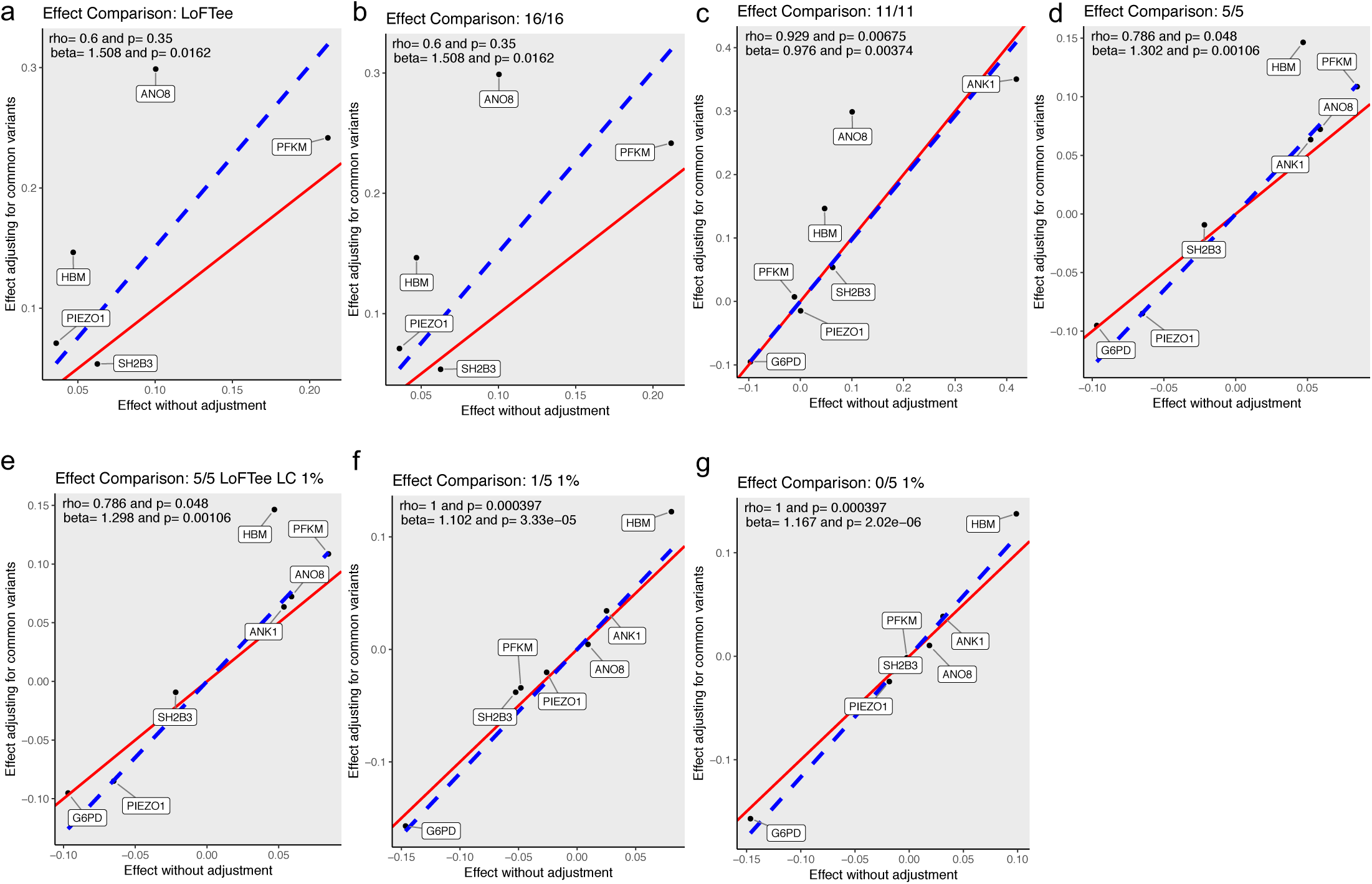
Impact of adjusting rare variant effects for common variants included in the polygenic score. Scatterplots indicate HbA1C gene-level effect sizes as estimated by burden tests with and without variants from the common variant PS included as covariates in the test. (**a-g**) Results are shown for each of the seven rare variant masks. We analyzed genes with nominal (*p*≤0.05) rare variant associations and within 125kb of a variant in the common variant PS. Results indicate that, on average, rare variant effects remain roughly the same when adjusting for common variants. Spearman’s rank correlation coefficients (*i.e.* rho) and associated *p*-values are indicated on plots. Red line indicates slope of 1. Slopes and *p*-values calculated for linear models are displayed on the plots; blue dotted lines indicate slopes estimated by the linear models.

**Supplementary Figure 6.**
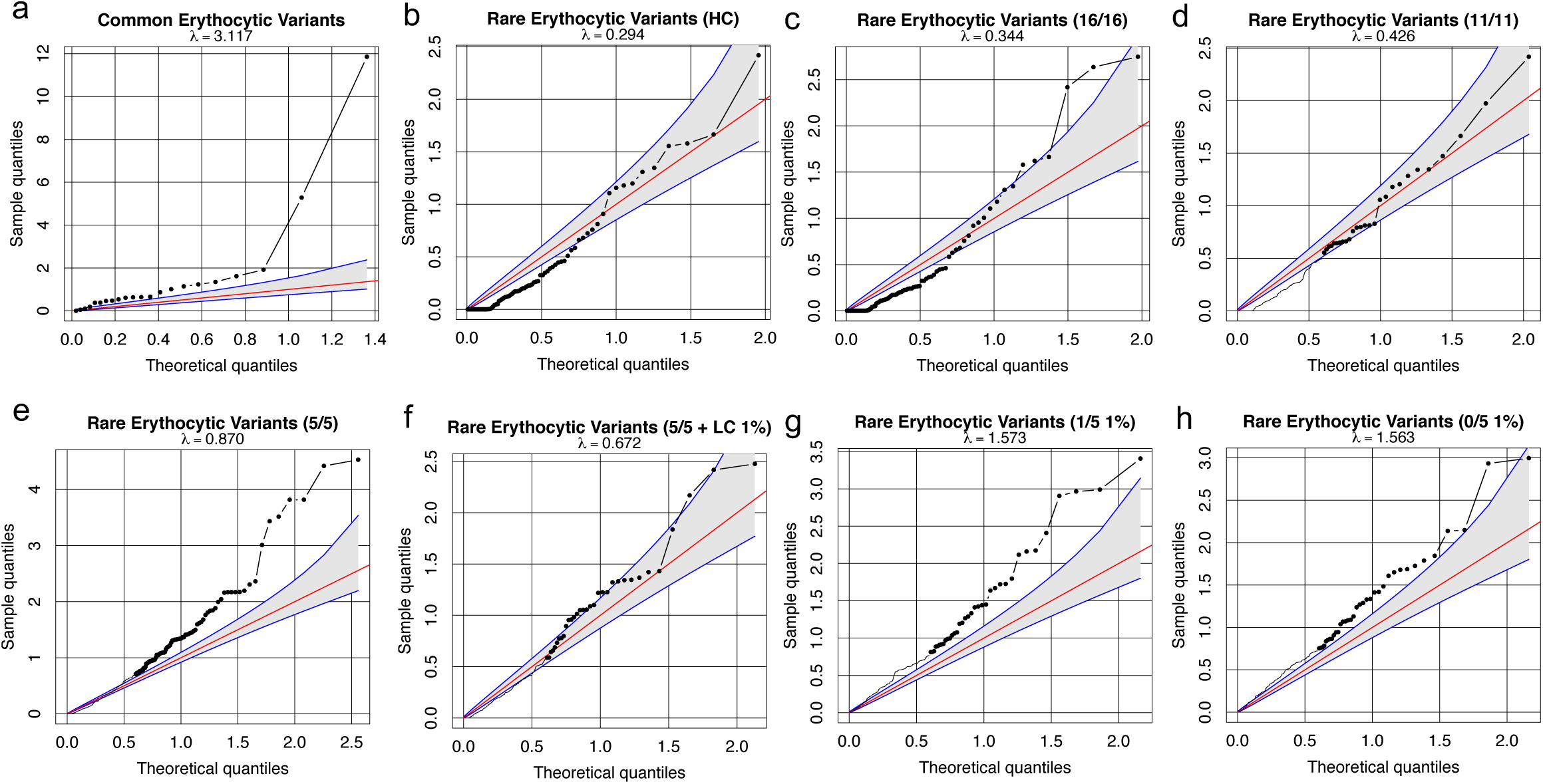
Testing for heterogeneity across ancestry for variants included in common variant and rare variant polygenic scores. We used a Cochran’s Q test to evaluate heterogeneity across ancestry-level single-variant and gene-level association results. QQ plots are shown for *p*-values from **(a)** single-variant Q tests for common variants and **(b-h)** gene-level Q tests for different rare variant masks (see ‘**Gene-level grouping**’ methods for details); included in each plot are the variants (or genes) included in the corresponding polygenic score. Departures above the diagonal red line suggest heterogeneity beyond the null expectation (blue lines indicate 90% confidence intervals for the null expectation), while lambda values indicate the ratio of the median observed chi square statistic to the median of the expected chi square statistic under the null; larger lambda values indicate larger deviations from the null. Abbreviations: ‘HC’ indicates the LoFTee mask; ‘5/5 + LC 1%’ indicates the “5/5 LoFTee LC 1%” mask.

## Supplementary Tables

**Supplementary Table 1. Study-level sample sizes.** Shown are the number of samples analyzed for each phenotype in the AMP-T2D-GENES and UKB datasets. Abbreviations: C-peptide-2hr: C-peptide level post 2-hour glucose tolerance test; C-peptide: C-peptide level; Insulin-2hr: insulin level post 2-hour glucose tolerance test; Glucose-2hr: glucose level post 2-hour glucose tolerance test; HOMAB: homeostatic model assessment of β-cell function; HOMAIR: homeostatic model assessment for insulin resistance; Insulin: fasting insulin; Glucose: fasting glucose; Hip-Circ: hip circumference; TG:HDL-C: Triglyceride-HDL-C ratio; Waist-Circ: waist circumference; RBC-count: red blood cell count; T2D: type 2 diabetes; HbA1C: hemoglobin A1C; Waist:Hip: waist-hip ratio; HDL-C high-density lipoprotein cholesterol; LDL-C: low-density lipoprotein cholesterol; TG: triglyceride; Total-C: total cholesterol; Systolic-BP: systolic blood pressure; Diastolic-BP: diastolic blood pressure; BMI: body mass index.

**Supplementary Table 2. Single variant associations from a meta-analysis of AMP-T2D-GENES and UKB.** All exome-wide significant (*p*≤1.8×10^-8^ [*i.e. p*≤4.3×10^-7^ with Bonferroni correction for 24 phenotypes]) coding variants and genome-wide significant (*p*≤2.1×10^-9^ [*i.e. p*≤5×10^-8^ with Bonferroni correction for 24 phenotypes]) non-coding variants are listed for each analyzed phenotype. Quantitative phenotype associations are for inverse normalized phenotype values. Genomic positions listed are based on GRCh37 genome assembly. Abbreviations: Allele: the effect allele (for which effect size is calculated) for the variant; Gene: the gene nearest to the variant; GWAS_locus: if variant listed is within 125kb of a known common variant significantly associated with the phenotype of interest, the gene nearest the most significantly variant (*i.e.* lowest *p*-value) within that GWAS locus is listed; Consequence: the consequence (Sequence Ontology term) of the variant as predicted by the Variant Effect Predictor (VEP); Impact: the impact of the variant as predicted by VEP; Min_freq: the lowest minor allele frequency across studies included in meta-analysis; Max_freq: the greatest minor allele frequency across studies included in meta-analysis.

**Supplementary Table 3. Single variant association summary statistics for untransformed HbA1C.** To calculate variant effect sizes on directly measured HbA1C, we computed single-variant associations for untransformed HbA1C following the same analysis strategy as for the inverse normalized phenotype values. We analyzed AMP-T2D-GENES and UKB samples separately and then meta-analyzed the results. All exome-wide significant (*p*≤4.3x10^-7^) coding variants and genome-wide significant (*p*≤5x10^-8^) non-coding variants from the meta-analysis are listed. Genomic positions listed are based on GRCh37 genome assembly. Abbreviations: Allele: the effect allele (for which effect size is calculated) for the variant; Gene: the gene nearest to the variant. GWAS_locus: if variant listed is within 125kb of a known significantly HbA1C associated common variant, the gene nearest the most significantly variant within that GWAS locus is listed; Consequence: the consequence (Sequence Ontology term) of the variant as predicted by the Variant Effect Predictor (VEP); Impact: the impact of the variant as predicted by VEP; Min_freq: the lowest minor allele frequency across studies included in meta-analysis; Max_freq: the greatest minor allele frequency across studies included in meta-analysis; Effect/St.Error: the estimated effect size of the effect allele and standard error of beta on untransformed HbA1C.

**Supplementary Table 4. Gene-level associations from a meta-analysis of AMP-T2D-GENES and UKB.** All exome-wide significant (*p*≤1.0×10^-7^ [*i.e. p*≤2.5×10^-6^ with Bonferroni correction for 24 phenotypes]) gene-level associations are listed. Results are from the minimum *p*-value consolidation method applied across results for each variant mask. Quantitative phenotype associations are for inverse normalized phenotype values. *P*-value/z-score are calculated via a sample-size weighted meta-analysis of burden test results.

**Supplementary Table 5. Gene-level associations for untransformed HbA1C.** We repeated our gene-level analysis but with untransformed HbA1C, rather than inverse normalized HbA1C, as an outcome variable. All mask-level results for genes with nominal (*p*≤0.05) gene-level associations from the inverse-variance weighted gene-level meta-analysis are listed. Abbreviations: Effect/St.Error: the estimated effect size calculated in a burden test using untransformed HbA1C as the outcome.

**Supplementary Table 6. Conditional *PIEZO1* analysis.** As the strongest associated (smallest *p*-value) *PIEZO1* burden test (5/5 variant mask; see ‘**Gene-level grouping**’ methods) with HbA1C explained a greater proportion of phenotype variance than a known^6^ common variant at the locus (rs837763), we evaluated potential LD between the rare variants and common variant by conducting *PIEZO1* burden tests both conditional (+) and not conditional (-) on the presence of rs837763. We calculated the proportion of variance explained (Variance Explained) by each resulting association. We calculated effect values within ancestry and combined them across ancestries with an inverse-weighted meta-analysis (see ‘**Proportion of variance explained calculations**’ method for details). Results suggest that inclusion of the common variant as a covariate has little impact on the proportion of variance explained by the *PIEZO1* rare variant burden test. Abbreviations: Effect: the beta value determined in the burden test; St.Error: the standard error of burden test beta value.

**Supplementary Table 7. Variants from the test sample included in rare variant polygenic scores.** Listed are all rare variants included in the rare variant polygenic scores that we evaluated (see ‘**Genes included in the rare variant polygenic score**’ methods for details). Variants shown are those in the AMP-T2D-GENES test sample; additional variants (not shown) in the UKB discovery sample were used to determine weights for the model. Genomic positions listed are based on GRCh37 genome assembly. Abbreviations: Allele: the effect allele (for which effect size is calculated) for the variant; Gene: the gene in which each variant is located.

**Supplementary Table 8. Performance of polygenic scores on T2D case reclassification.** For the best performing rare variant polygenic score (*i.e.* ‘loose, nested’) model and a previously published common variant polygenic score, we calculated the number of cases (Cases reclassed) and controls (Controls reclassed) who had an unadjusted HbA1C value below 6.5% but whose T2D status was “reclassified” based on an adjusted HbA1C value above 6.5%. We then calculated the number of cases (Matched-Cases) and controls (Matched-Controls) among a random sample of individuals matched on unadjusted HbA1C levels, cohort, and ancestry. The OR and *p*-value indicate the enrichment of cases (according to a Fisher’s Exact test) in the analysis based on the polygenic score relative to the analysis based on the random sample. Results are shown separately for each ancestry and from a meta-analysis across all (All) ancestries.

**Supplementary Table 9. Mean number of alleles carried by individuals in each polygenic score.** For the best performing rare variant polygenic score (*i.e.* ‘loose, nested’) model and the common variant polygenic score, shown are the total number of variants included in the score and the distribution of variants (Mean, St.Error, 95% confidence interval) carried by each individual. Results are stratified by individual ancestry.

**Supplementary Table 10. Effect sizes of rare and common variant associations used for polygenic scores.** The number of unique genes or variants, as well as their mean absolute effect as determined in the discovery sample (*i.e*. UKB for gene-level rare variant analysis; Wheeler et al.^6^ for common variants), is reported for: (a) all genes in the best performing rare variant polygenic score (‘loose, nested’ model) and (b) all variants in the common variant polygenic score. Effect sizes are shown in untransformed HbA1C units (*i.e.* percent glycated hemoglobin). Abbreviations: Genes/Loci: number of unique genes (rare variant model) or loci (common variant model) included in the polygenic score. Mean(|Effect|) and St.Error(|effect|): mean and standard error of absolute effect in untransformed HbA1C units; 95% of mean(|effect|): 95% confidence interval of mean absolute effect.

**Supplementary Table 11. Mean sample adjustments for rare and common variant models.** Shown are the estimated mean absolute HbA1C adjustments (Mean[|adjustment|], SE[|adjustment|], 95% CI of mean[|adjustment|]), as determined in the test sample (*i.e.* AMP-T2D-GENES), for (a) the best performing rare variant polygenic score (‘loose, nested’ model) across all samples (All) as well as within each ancestry and (b) the common variant polygenic score within each ancestry. Effect sizes are shown in untransformed HbA1C units (*i.e.* percent glycated hemoglobin).

**Supplementary Table 12. AMP-T2D-GENES cohort-level phenotype statistics.** For each cohort within the AMP-T2D-GENES study and each analyzed phenotype, shown are the number of samples, average phenotype values, average age, and sex distribution. Standard deviations are shown in parentheses. Phenotype value units are labeled in table header.

**Supplementary Table 13. Single variant associations in AMP-T2D-GENES samples.** All exome-wide significant (*p*≤4.3x10^-7^) coding variants and genome-wide significant (*p*≤5x10^-8^) non-coding variants are listed for each analyzed phenotype in AMP-T2D-GENES. We inverse normalized quantitative phenotype values prior to analysis. Abbreviations: Allele: the effect allele (for which effect size is calculated) for the variant; Gene: the gene nearest to the variant; GWAS_locus: if variant listed is within 125kb of a known significantly associated common variant, the gene nearest the most significantly variant within that GWAS locus is listed; Consequence: the consequence (Sequence Ontology term) of the variant as predicted by the Variant Effect Predictor (VEP); Impact: the impact of the variant as predicted by VEP; MAF: minor allele frequency; Effect: the estimated effect size of the effect allele on the inverse normalized phenotype value.

**Supplementary Table 14. Gene-level associations in AMP-T2D-GENES samples.** All exome-wide significant (*p*≤2.5x10^-6^) gene-level signals are listed for each gene, phenotype, and mask. We calculated betas and *p*-values with a burden test using inverse normalized phenotype values. Abbreviations: N: number of samples analyzed. Variants: number of variants included in burden test. Fraction_rare: Fraction of individuals analyzed carrying rare variants included in the burden test.

**Supplementary Table 15. UKB phenotype statistics.** For each analyzed phenotype, shown are the sample size, distribution of phenotype values, average age, and sex composition of UKB samples in the analysis. Parentheses indicate standard deviations. Phenotype value units are labeled in table header.

**Supplementary Table 16. UKB quality control metrics.** We calculated a variety of metrics for each sample in the UKB (see ‘**UK Biobank sample-level quality control**’ methods) and excluded samples from analysis that were outliers according to one or more metrics. Shown are the number of samples removed by each metric, as well as the total number of samples removed. Abbreviations: call_rate: fraction of variants with called genotypes; het: inbreeding coefficient; het_high: inbreeding coefficient for variants with MAF ≤0.03; het_low: inbreeding coefficient for variants with MAF <0.03; n_called: number of homozygous reference alleles + number of heterozygous variants + number of homozygous variants; n_het: number of heterozygous variants; n_hom_var: number of homozygous alternate variants; n_non_ref: number of heterozygous + number of homozygous variants; r_het_hom_var: homozygous variant ratio across all variants; r_ti_tv: transition/transversion; PCA: principal component analysis of ancestry.

**Supplementary Table 17. Single variant associations in UKB samples.** All exome-wide significant (*p* 4.3x10^-7^) coding variants and genome-wide significant (*p*≤5x10^-8^) non-coding variants are listed for each analyzed phenotype. We inverse normalized quantitative phenotype values prior to analysis. Abbreviations: Allele: the effect allele (for which effect size is calculated) for the variant. Gene: the gene nearest to the variant. GWAS_locus: the gene nearest to the variant if a GWAS signal lies nearby. Consequence: the consequence (Sequence Ontology term) of the variant as predicted by the Variant Effect Predictor (VEP). Impact: the impact of the variant as predicted by VEP. MAF: minor allele frequency. Effect: the estimated effect size of the effect allele on the inverse normalized phenotype value.

**Supplementary Table 18. Gene-level associations in UKB samples.** All exome-wide significant (*p*≤2.5x10^-6^) gene-level associations are listed for each phenotype and each variant mask. Quantitative phenotype associations are for inverse normalized phenotype values. Effects/*P*-values are calculated with a burden test. Abbreviations: N: number of samples analyzed. Variants: number of variants included in burden test. Fraction_rare: Fraction of individuals analyzed carrying rare variants included in the burden test.

**Supplementary Table 19. Mask-level associations from the AMP-T2D-GENES and UKB meta-analysis.** All exome-wide significant (*p*≤2.5x10^-6^) gene-level signals are listed for each phenotype and variant mask. *P*-values/z-scores are calculated via a sample size weighted meta-analysis.

**Supplementary Table 20. Performance of the rare variant polygenic score with and without *COG8*.** We repeated the analysis in **Supplementary Table 8** after removing variants in *COG8* from the rare variant polygenic score. The table shows the results of the original analysis with COG8 (+) as well as the new analysis without it (-).

**Supplementary Table 21. Performance of the common variant polygenic score with and without rs1050828.** We repeated the analysis in **Supplementary Table 8** after removing rs1050828 from the common variant polygenic score. The table shows the results of the original analysis with rs1050828 (+) as well as the new analysis without it (-).

## Acknowledgements

This project was supported by R01DK125490 and UM1DK105554. JBC is supported by a NIDDK Pathway to Independence Award (K99DK127196). AL was supported by Grant 2020096 from the Doris Duke Charitable Foundation. JBM was supported by NIH R01 DK078616 and R01HL151855. JCF was supported by NHLBI K24 HL157960. JIR was supported by the National Center for Advancing Translational Sciences, CTSI grant UL1TR001881, and the National Institute of Diabetes and Digestive and Kidney Disease Diabetes Research Center (DRC) grant DK063491 to the Southern California Diabetes Endocrinology Research Center. Infrastructure for the CHARGE Consortium is supported in part by the National Heart, Lung, and Blood Institute (NHLBI) grant R01HL105756. Also supported in part by the National Institutes of Health, National Heart, Lung, Long and Blood Institute (NHLBI) contract 1R01HL151855 and the National Institute of Diabetes and Digestive and Kidney Diseases contract UM1DK078616. MSU was supported by K23DK114551.

## Notes

### Competing Interest Statement

The authors have declared no competing interest.

### Author Declarations

This study involves only openly available human data, which can be obtained from (a) the NCBI database of genotypes and phenotypes (dbGaP) or (b) through the UK Biobank.

