## Supplementary material for "A combined polygenic score of 21,293 rare and 22 common variants significantly improves diabetes diagnosis based on hemoglobin A1C levels": AMP-T2D-GENES Consortia Authors

**AMP-T2D-GENES Consortium**

Carlos A. Aguilar-Salinas^1^, Gil Atzmon^2,3,4^, Francisco Barajas-Olmos^5^, Nir Barzilai^2,4^, John Blangero^6^, Eric Boerwinkle^7,8^, Lori L. Bonnycastle^9^, Erwin Bottinger^10^, Donald W Bowden^11,12,13^, Federico Centeno- Cruz^5^, John C. Chambers^14,15^, Edmund Chan^16^, Juliana Chan^17,18,19,20^, Ching-Yu Cheng^21,22,23^, Yoon Shin Cho^24^, Cecilia Contreras-Cubas^5^, Emilio Córdova^5^, Adolfo Correa^25^, Ralph A. DeFronzo^26^, Ravindranath Duggirala^6^, Josée Dupuis^27^, Humberto García-Ortiz^5^, Christian Gieger^28,29,30^, Benjamin Glaser^31^, Clicerio Gonzalez^32^, Ma Elena Gonzalez^33^, Niels Grarup^34^, Leif Groop^35,36^, Myron Gross^37^, Christopher Haiman^38^, Sohee Han^39^, Craig L. Hanis^7^, Torben Hansen^34^, Nancy L. Heard-Costa^40,41^, Brian E. Henderson^38^, Juan Manuel Malacara Hernandez^42^, Mi Yeong Hwang^39^, Sergio Islas-Andrade^5^, Marit E. Jørgensen^43,44,45^, Hyun Min Kang^46^, Bong-Jo Kim^39^, Young Jin Kim^39^, Heikki A. Koistinen^47,48,49^, Jaspal Singh Kooner^50,51,52^, Johanna Kuusisto^53^, Soo Heon Kwak^54^, Markku Laakso^53^, Leslie A. Lange^55^, Juyoung Lee^39^, Jong-Young Lee^56^, Donna M. Lehman^26^, Allan Linneberg^57,58,59^, Jianjun Liu^16,60,61^, Ruth Loos^10,62^, Valeriya Lyssenko^33,63^, Ronald C.W. Ma^17,18,19,20^, Angélica Martínez-Hernández^5^, James B. Meigs^64,65,66^, Thomas Meitinger^67,68^, Elvia Mendoza- Caamal^5^, Karen L. Mohlke^69^, Andrew D. Morris^70,71^, Alanna C. Morrison^7,72^, Maggie C.Y. Ng^11,12,13^, Peter Nilsson^73^, Christopher J. O’Donnell^74,75,76,77^, Lorena Orozco^5^, Colin N.A. Palmer^78^, Kyong Soo Park^54,79,80^, Oluf Pedersen^34^, Wendy S. Post^81^, Michael Preuss^10^, Bruce M. Psaty^82,83^, Ramachandran S. Vasan^40,84^, Alexander P. Reiner^85^, Cristina Revilla-Monsalve^5^, Stephen S. Rich^86^, Jerome I. Rotter^87^, Zhe Wang^10^, Maria Eugenia Garay Sevilla^88,89^, Xueling Sim^60^, Rob Sladek^90,91,92^, Kerrin S. Small^93^, Wing Yee So^17,18,19^, Timothy D. Spector^93^, Konstantin Strauch^29,94^, Tim M. Strom^67,95^, E-Shyong Tai^16,60,96^, Claudia H.T. Tam^17,18,19^, Yik Ying Teo^60,96^, Farook Thameem^97^, Brain Tomlinson^98^, Russell P. Tracy^99,100^, Tiinamaija Tuomi^101,102^, Jaakko Tuomilehto^103,104,105,106^, Teresa Tusié-Luna^107,108^, Rob M. van Dam^16,60,109^, James G. Wilson^110^, Daniel R. Witte^111,112^, Tien-Yin Wong^21,22,23^, Peter Dornbos^64,74,113^, Jason A. Flannick^64,74,113^, Jose C. Florez^64,65,114^, Mark I. McCarthy^115^, Michael Boehnke^46^, Noël P. Burtt^64^

**Affiliations:**

^1^Instituto Nacional de Ciencias Medicas y Nutricion, Mexico City, Mexico

^2^Department of Medicine, Albert Einstein College of Medicine, New York, NY, USA. ^3^Faculty of Natural Science, University of Haifa, Haifa, Israel.

^4^Department of Genetics, Albert Einstein College of Medicine, New York, NY, USA

^5^Instituto Nacional de Medicina Genómica, Mexico City, Mexico

^6^Department of Human Genetics and South Texas Diabetes and Obesity Institute, University of Texas Rio Grande Valley, Brownsville and Edinburg, TX, USA

^7^Human Genetics Center, School of Public Health, The University of Texas Health Science Center at Houston, Houston, TX, USA

^8^Human Genome Sequencing Center, Baylor College of Medicine, Houston, TX, USA.

^9^Medical Genomics and Metabolic Genetics Branch, National Human Genome Research Institute, National Institutes of Health, Bethesda, MD, USA

^10^The Charles Bronfman Institute for Personalized Medicine, Icahn School of Medicine at Mount Sinai, New York, NY, USA

^11^Center for Diabetes Research, Wake Forest School of Medicine, Winston-Salem, NC, USA.

^12^Center for Genomics and Personalized Medicine Research, Wake Forest School of Medicine, Winston-Salem, NC, USA.

^13^Department of Biochemistry, Wake Forest School of Medicine, Winston-Salem, NC, USA.

^14^Department of Epidemiology and Biostatistics, Imperial College London, London, UK.

^15^Lee Kong Chian School of Medicine, Nanyang Technological University, Singapore, Singapore.

^16^Department of Medicine, Yong Loo Lin School of Medicine, National University of Singapore and National University Health System, Singapore, Singapore.

^17^Department of Medicine and Therapeutics, The Chinese University of Hong Kong, Hong Kong, China.

^18^Chinese University of Hong Kong-Shanghai Jiao Tong University Joint Research Centre in Diabetes Genomics and Precision Medicine, The Chinese University of Hong Kong, Hong Kong, China.

^19^Hong Kong Institute of Diabetes and Obesity, The Chinese University of Hong Kong, Hong Kong, China.

^20^Li Ka Shing Institute of Health Sciences, The Chinese University of Hong Kong, Hong Kong, China.

^21^Singapore Eye Research Institute, Singapore National Eye Centre, Singapore, Singapore

^22^Ophthalmology & Visual Sciences Academic Clinical Program (Eye ACP), Duke-NUS Medical School, Singapore, Singapore

^23^Department of Ophthalmology, Yong Loo Lin School of Medicine, National University of Singapore and National University Health System, Singapore, Singapore.

^24^Department of Biomedical Science, Hallym University, Chuncheon, South Korea

^25^Department of Medicine, University of Mississippi Medical Center, Jackson, MS, USA.

^26^Department of Medicine, University of Texas Health San Antonio (aka University of Texas Health Science Center at San Antonio), San Antonio, TX, USA

^27^Department of Biostatistics, Boston University School of Public Health, Boston, MA, USA.

^28^Research Unit Molecular Epidemiology, Helmholtz Zentrum München, German Research Center for Environmental Health, Neuherberg, Germany.

^29^Institute of Epidemiology, Helmholtz Zentrum München, German Research Center for Environmental Health, Neuherberg, Germany.

^30^German Center for Diabetes Research (DZD), Neuherberg, Germany.

^31^Endocrinology and Metabolism Service, Hadassah-Hebrew University Medical Center, Jerusalem, Israel.

^32^Unidad de Investigacion en Diabetes y Riesgo Cardiovascular, Instituto Nacional de Salud Publica, Cuernavaca, Mexico.

^33^Centro de Estudios en Diabetes, Mexico City, Mexico.

^34^Novo Nordisk Foundation Center for Basic Metabolic Research, Faculty of Health and Medical Sciences, University of Copenhagen, Copenhagen, Denmark.

^35^Department of Clinical Sciences, Diabetes and Endocrinology, Lund University Diabetes Centre, Malmö, Sweden.

^36^Institute for Molecular Medicine Finland, University of Helsinki, Helsinki, Finland.

^37^Department of Laboratory Medicine and Pathology, University of Minnesota, Minneapolis, MN, USA.

^38^Department of Preventive Medicine, Keck School of Medicine of USC, Los Angeles, CA, USA.

^39^Division of Genome Research, Center for Genome Science, National Institute of Health, Chungcheongbuk-do, South Korea.

^40^Boston University and National Heart Lung and Blood Institute’s Framingham Heart Study, Framingham, MA, USA.

^41^Department of Neurology, Boston University School of Medicine, Boston, MA, USA.

^42^Department of Medical Science, División of Health Science. University of Guanjuato. Campus León. León, Gto. México.

^43^Steno Diabetes Center Copenhagen, Gentofte, Denmark.

^44^National Institute of Public Health, University of Southern Denmark, Copenhagen, Denmark.

^45^Greenland Centre for Health Research, University of Greenland, Nuuk, Greenland.

^46^Department of Biostatistics and Center for Statistical Genetics, University of Michigan, Ann Arbor, MI, USA.

^47^Department of Public Health Solutions, Finnish Institute for Health and Welfare, Helsinki, Finland.

^48^University of Helsinki and Department of Medicine, Helsinki University Central Hospital, Helsinki, Finland.

^49^Minerva Foundation Institute for Medical Research, Helsinki, Finland.

^50^Department of Cardiology, Ealing Hospital, London North West Healthcare NHS Trust, London, UK.

^51^MRC-PHE Centre for Environment and Health, Imperial College London, London, UK. Imperial College Healthcare NHS Trust, Imperial College London, London, UK. ^52^National Heart and Lung Institute, Imperial College London, London, UK.

^53^Institute of Clinical Medicine, Internal Medicine, University of Eastern Finland and Kuopio University Hospital, Kuopio, Finland.

^54^Department of Internal Medicine, Seoul National University Hospital, Seoul, South Korea.

^55^Department of Medicine, University of Colorado Denver, Anschutz Medical Campus, Aurora, CO, USA.

^56^Oneomics Soonchunhyang Mirae Medical Center, Bucheon-si Gyeonggi-do, Republic of Korea.

^57^Department of Clinical Medicine, Faculty of Health and Medical Sciences, University of Copenhagen, Copenhagen, Denmark.

^58^Center for Clinical Research and Prevention, Bispebjerg and Frederiksberg Hospital, Copenhagen, Denmark.

^59^Department of Clinical Experimental Research, Rigshospitalet, Copenhagen, Denmark.

^60^Saw Swee Hock School of Public Health, National University of Singapore and National University Health System, Singapore, Singapore.

^61^Genome Institute of Singapore, Agency for Science Technology and Research, Singapore, Singapore.

^62^The Mindich Child Health and Development Institute, Ichan School of Medicine at Mount Sinai, New York, NY, USA.

^63^Department of Clinical Science, University of Bergen, Bergen, Norway.

^64^Program in Medical and Population Genetics, Broad Institute of MIT and Harvard, Cambridge, MA, USA.

^65^Department of Medicine, Harvard Medical School, Boston, MA, USA.

^66^Division of General Internal Medicine, Massachusetts General Hospital, Boston, MA, USA.

^67^Institute of Human Genetics, Technical University of Munich, Munich, Germany.

^68^German Centre for Cardiovascular Research (DZHK), Partner Site Munich Heart Alliance, Munich, Germany.

^69^Department of Genetics, University of North Carolina Chapel Hill, Chapel Hill, NC, USA.

^70^Wellcome Centre for Human Genetics, Nuffield Department of Medicine, University of Oxford, Oxford, UK.

^71^Department of Biostatistics, University of Liverpool, Liverpool, UK.

^72^Department of Epidemiology, Human Genetics, and Environmental Sciences, School of Public Health, The University of Texas Health Science Center at Houston, Houston, TX, USA.

^73^Department of Clinical Sciences, Medicine, Lund University, Malmö, Sweden.

^74^Department of Pediatrics, Harvard Medical School, Boston, MA, USA.

^75^Section of Cardiology, Department of Medicine, VA Boston Healthcare, Boston, MA, USA.

^76^Brigham and Women’s‚ Hospital, Boston, MA, USA.

^77^Intramural Administration Management Branch, National Heart Lung and Blood Institute, NIH, Framingham, MA, USA.

^78^Pat Macpherson Centre for Pharmacogenetics and Pharmacogenomics, University of Dundee, Dundee, UK

^79^Department of Internal Medicine, Seoul National University College of Medicine, Seoul, South Korea.

^80^Department of Molecular Medicine and Biopharmaceutical Sciences, Graduate School of Convergence Science and Technology, Seoul National University, Seoul, South Korea.

^81^Division of Cardiology, Department of Medicine, Johns Hopkins University, Baltimore, MD, USA.

^82^Cardiovascular Health Research Unit, Departments of Medicine, Epidemiology, and Health Services, University of Washington, Seattle, WA, USA.

^83^Kaiser Permanente Washington Research Institute, Seattle, WA, USA.

^84^Preventive Medicine & Epidemiology, and Cardiovascular Medicine, Medicine, Boston University School of Medicine, and Epidemiology, Boston University School of Public health, Boston, MA, USA.

^85^Fred Hutchinson Cancer Research Center, Seattle, WA, USA.

^86^Center for Public Health Genomics, University of Virginia School of Medicine, Charlottesville, VA, USA

^87^The Institute for Translational Genomics and Population Sciences, Department of Pediatrics, The Lundquist Institute for Biomedical Innovation (formerly Los Angeles Biomedical Research Institute) at Harbor-UCLA Medical Center, Torrance, CA, USA.

^88^Digital Health Center, Hasso Plattner Institute, University of Potsdam, Prof.-Dr.-Helmert-Str. 2-3, Potsdam, Germany.

^89^Hasso Plattner Institute for Digital Health at Mount Sinai, Icahn School of Medicine at Mount Sinai, One Gustave L. Levy Place, New York, NY, USA.

^90^Department of Human Genetics, McGill University, Montreal, QC, Canada.

^91^Division of Endocrinology and Metabolism, Department of Medicine, McGill University, Montreal, QC, Canada.

^92^McGill University and Génome Québec Innovation Centre, Montreal, QC, Canada.

^93^Department of Twin Research and Genetic Epidemiology, King’s College London, London, UK.

^94^Institute for Medical Informatics Biometry and Epidemiology, Ludwig-Maximilians University, Munich, Germany.

^95^Institute of Human Genetics, Helmholtz Zentrum München, German Research Center for Environmental Health, Neuherberg, Germany.

^96^Life Sciences Institute, National University of Singapore, Singapore, Singapore.

^97^Department of Biochemistry, Faculty of Medicine, Health Science Center, Kuwait University, Safat, Kuwait.

^98^Faculty of Medicine, Macau University of Science & Technology, Macau, China.

^99^Department of Pathology and Laboratory Medicine, The Robert Larner M.D. College of Medicine, University of Vermont, Burlington, VT, USA.

^100^Department of Biochemistry, The Robert Larner M.D. College of Medicine, University of Vermont, Burlington, VT, USA.

^101^Department of Endocrinology, Abdominal Centre, Helsinki University Hospital, Helsinki, Finland; Institute for Molecular Medicine Finland, University of Helsinki, Helsinki, Finland

^102^Folkhaalsan Research Centre, Helsinki, Finland; Research Programs Unit, Clinical and Molecular Medicine, University of Helsinki, Helsinki, Finland.

^103^Diabetes Prevention Unit, National Institute for Health and Welfare, Helsinki, Finland.

^104^Center for Vascular Prevention, Danube University Krems, Krems, Austria.

^105^Diabetes Research Group, King Abdulaziz University, Jeddah, Saudi Arabia.

^106^Instituto de Investigacion Sanitaria del Hospital Universario LaPaz (IdiPAZ), University Hospital LaPaz, Autonomous University of Madrid, Madrid, Spain.

^107^Unidad de Biología Molecular y Medicina Genómica, Instituto Nacional de Ciencias Médicas y Nutrición Salvador Zubirán, Mexico City, Mexico.

^108^Departamento de Medicina Genómica y Toxiología Ambiental, Instituto de Investigaciones Biomédicas, UNAM, Mexico City, Mexico.

^109^Department of Nutrition, Harvard T.H. Chan School of Public Health, Boston, MA, USA.

^110^Department of Physiology and Biophysics, University of Mississippi Medical Center, Jackson, MS, USA.

^111^Department of Public Health, Aarhus University, Aarhus, Denmark.

^112^Danish Diabetes Academy, Odense, Denmark.

^113^Boston Children‚Äôs Hospital, Boston, Massachusetts, USA.

^114^Diabetes Unit and Center for Genomic Medicine, Massachusetts General Hospital, Boston, MA, USA.

^115^Genentech, South San Francisco, California, USA.
